# Identifying interstitial lung disease with large language models on clinical notes and claims

**DOI:** 10.64898/2026.10.30.26366942

**Authors:** Guillaume Larghero, Sebastian Schneeweiss, Bowen Gu, Priyanka Anand, Kueiyu Joshua Lin, Jie Yang

**Affiliations:** Division of Pharmacoepidemiology and Pharmacoeconomics, Department of Medicine, Mass General Brigham, Boston, MA, USA; Harvard Medical School, Boston, MA, USA; Department of Biomedical Informatics, Harvard Medical School, Boston, MA, USA; Kempner Institute for the Study of Natural and Artificial Intelligence, Harvard University, Cambridge, MA, USA; Broad Institute of MIT and Harvard, Cambridge, MA, USA

**Keywords:** Interstitial lung disease, Pharmacovigilance, Electronic health record, Administrative claims, Large language models, Immune checkpoint inhibitors, Antibody-drug conjugates

## Abstract

**Background:** Interstitial lung disease (ILD) is a potentially fatal toxicity of immune check-point inhibitors and antibody-drug conjugates. Drug-safety studies in healthcare databases identify ILD with claims-based algorithms that trade positive predictive value (PPV) against sensitivity. We assessed whether large language models (LLMs) improve on these algorithms, with and without electronic health record (EHR) notes, and whether these gains translate into measurable epidemiological benefits.

**Methods:** In 309 oncology patients with ILD diagnosis codes at Mass General Brigham, adjudicated by two pulmonologists, we compared 12 open-access LLMs with 20 published claims-based algorithms. LLMs received claims, EHR notes, or both, without task-specific training. Measures were PPV and yield (proportion classified positive). We projected the attenuation of a drug-induced ILD odds ratio under non-differential outcome misclassification.

**Findings:** LLMs extended the PPV–yield frontier of the published algorithms with all three inputs. Across the 36 model–input evaluations, LLM PPV exceeded that of the best algorithm without specialist-coding requirements (Algorithm 4) by 14.3 percentage points on average. With claims alone, Qwen3-8B achieved 89.8% PPV (95% CI 78.2–95.6) at 15.9% yield, versus 86.7% at 4.9% for the highest-PPV algorithm. With notes alone, Qwen3-32B achieved 78.9% PPV at 49.2% yield, versus 64.6% at 51.1% for Algorithm 4. With combined claims and notes, Qwen3-14B achieved 100.0% PPV at 24.3% yield. Under a simulated drug-safety scenario with a true odds ratio of 3.0, the projected observed odds ratio was 2.67 with combined inputs versus 1.07 with Algorithm 4, corresponding to a 46-fold difference in the estimated sample size required for 80% power (591 versus 27,200 patients).

**Interpretation:** Zero-shot LLMs offer a higher-PPV alternative to rule-based claims pheno-typing for ILD, on all three input types. Because the claims-only configuration uses the same claims data as existing algorithms, it could be deployed in administrative databases without linked notes, and may recover drug-safety signal otherwise lost to outcome misclassification.

**Funding:** National Institute on Aging, National Library of Medicine, PCORI, and the National Institute of Arthritis and Musculoskeletal and Skin Diseases.

**Research in context:** *Evidence before this study:* We searched PubMed up to August 2026 for studies on claims or electronic health record (EHR)-based identification of interstitial lung disease and on machine-learning or large language model (LLM) phenotyping of clinical conditions. Search terms included “interstitial lung disease or pneumonitis”, “administrative claims”, “computational phenotyping”, “rule-based algorithms”, “machine learning”, and “large language models”. We also screened the reference lists of relevant articles. The literature search suggested that identifying ILD in longitudinal claims data is unreliable because the relevant diagnosis codes are nonspecific, and validated rule-based algorithms face a structural tradeoff between PPV and coverage. Supervised machine-learning classifiers have outperformed rule-based ILD identification on electronic health record data, and zero-shot LLMs have already shown the ability to phenotype chronic conditions from structured codes. No study, however, had compared a zero-shot LLM head-to-head against validated, published claims-based algorithms for a specific, high-stakes drug-safety outcome on a single fixed cohort, isolated the contribution of the input substrate from that of the model, or quantified the downstream consequence for pharmacoepidemiologic effect estimation.

*Added value of this study:* On a single, fixed, expert-adjudicated cohort, the evaluated LLMs achieved higher positive predictive value (PPV) than all 20 published claims-based algorithms at comparable yield (the proportion of patients classified as ILD-positive), whether the models received structured claims, clinical notes, or both. Across the 36 model–input evaluations, PPV exceeded that of the most inclusive published rule by 19.7 percentage points on average, and did so in 33 of 36 evaluations. We show that clinical notes generally increased yield, while the resulting PPV–yield trade-off depended on the model and input combination. Restricting the input to a single EHR note type preserved the advantage over rule-based phenotyping in matched comparisons on the patients possessing that EHR note type. Most consequentially, this improvement reduces the sample size required to detect a hypothesised drug-ILD association from approximately 27,200 patients for the best existing algorithm for the task to 591 when using LLMs, and offer a deployable upgrade for the note-free administrative databases where rule-based algorithms are the current standard.

*Implications of all the available evidence:* For pharmacovigilance of treatment-related ILD, the limiting step is often not the size of the available claims database but the misclassification that erodes the very signal under surveillance. Replacing rule-based claims phenotyping algorithms with zero-shot LLM-based alternatives transforms a high-PPV, low-yield classifier into one that recovers many more patients at the same PPV, converting a classifier that can render drug-safety associations statistically invisible into one that makes them detectable in cohorts of feasible size. Pending external validation, this offers a practical route to more sensitive post-marketing surveillance of the pulmonary toxicity of immune checkpoint inhibitors and antibody-drug conjugates. More broadly, we hypothesise that this advantage is not specific to ILD: zero-shot LLM phenotyping may recover drug-safety signal for other outcomes that diagnosis codes capture poorly but that are well documented in clinical text, a possibility that warrants dedicated evaluation.

## 1 Introduction

Drug-induced interstitial lung disease (ILD) is a recognized class effect of immune checkpoint inhibitors (ICIs) and antibody-drug conjugates (ADCs), and one of the most consequential toxicities of modern oncologic therapy.^1,2^ In a pooled analysis of nine trastuzumab deruxtecan trials, 15.4% of patients developed adjudicated drug-related ILD or pneumonitis, including 2.2% grade 5 events,^3^ and pneumonitis is the most common cause of death attributed to anti-PD-1/PD-L1 therapy.^4,5^ As these agents are used more widely, drug-safety studies increasingly rely on large claims databases, in which confirming cases by manual chart review does not scale. ^6^

Automated identification relies instead on rules built from diagnosis codes, and for ILD these codes are nonspecific. ^7–9^ J18.9 (pneumonia, unspecified organism), for example, is recorded for both ILD and infectious pneumonia. ^6,7^ Rule-based algorithms therefore trade positive predictive value (PPV) against the number of cases identified. ^8,9^ Across 20 claims-based algorithms evaluated by Shen and colleagues, whose cohort is the benchmark for the present study, broadly inclusive rules reached a PPV near 59%, whereas rules restricted to specialist-coded events reached PPVs in the high 80s but identified only a small fraction of patients. ^10^ Both failures matter for drug-safety studies: false-positive outcomes attenuate exposure-outcome associations, and overly restrictive definitions leave too few cases for adequately powered analyses. ^11–13^

The evidence on which an ILD diagnosis rests, such as imaging, pulmonary function testing and specialist assessment, is recorded in clinical notes rather than in claims, but it is unstructured and voluminous. ^14^ Large language models have shown the ability to interpret such unstructured information at scale, in both clinical notes and claims.^15–17^ Unlike a fixed rule, an LLM can weigh whether an ILD code is corroborated or contradicted by other evidence in the record. Zero-shot LLMs have already outperformed rule-based methods for phenotyping chronic conditions from structured codes, ^18,19^ and have supported case adjudication in observational research.^20^ For ILD specifically, supervised machine-learning classifiers have outperformed rule-based identification on EHR data. ^6^ No study, however, has compared LLMs with validated claims-based algorithms for a drug-safety outcome on the same patients, separated the contribution of the input data from that of the model, or asked whether better phenotyping changes the resulting effect estimates. ^19,20^

We sought to evaluate whether LLMs could improve ILD ascertainment for pharmacoepidemiologic drug-safety studies beyond investigator-defined algorithms, and whether any improvement would change the resulting drug–outcome effect estimates.

## 2 Methods

### Study design and reference standard

We conducted a benchmarking study using an expert-adjudicated cohort of 360 oncology patients from Mass General Brigham (MGB), the largest healthcare system in Massachusetts, USA, between January 1, 2017, and December 31, 2024. ^10^ Patients were required to have at least two International Classification of Diseases, Tenth Revision, Clinical Modification (ICD-10-CM) ILD diagnosis codes (J84.113, J84.114, J84.89, J84.9, J70.2, J70.3, J70.4, J70.8, J70.9, or J18.9) recorded 7–180 days apart and a diagnosis of breast, lung, colorectal, or gynecologic cancer. Patients were aged ≥ 18 years, or ≥ 66 years for Medicare beneficiaries, and had at least 365 days of continuous enrollment before the cohort entry date (CED). The CED was defined as the date of the second qualifying ILD code. Two pulmonologists independently used a standardized abstraction tool to review each patient’s full EHR and assigned Probable ILD, Possible ILD, Not ILD, or Insufficient Evidence (inter-rater *κ* = 0.87). For binary evaluation, Probable and Possible ILD were combined into the positive class, following the source study.^10^

We excluded 43 patients classified as Insufficient Evidence and 8 without retrievable EHR data, leaving a cohort of 309 patients with an adjudicated label and at least one clinical note within the observation window (Supplementary Fig. S1). Figure 1 summarises the study cohort, reference algorithms, and LLM evaluation workflow. We evaluated three primary input configurations: *Claims*, comprising structured claims alone; *EHR notes*, comprising clinical notes from HER alone; and *Fused*, comprising claims and clinical notes together.

**Figure 1.**
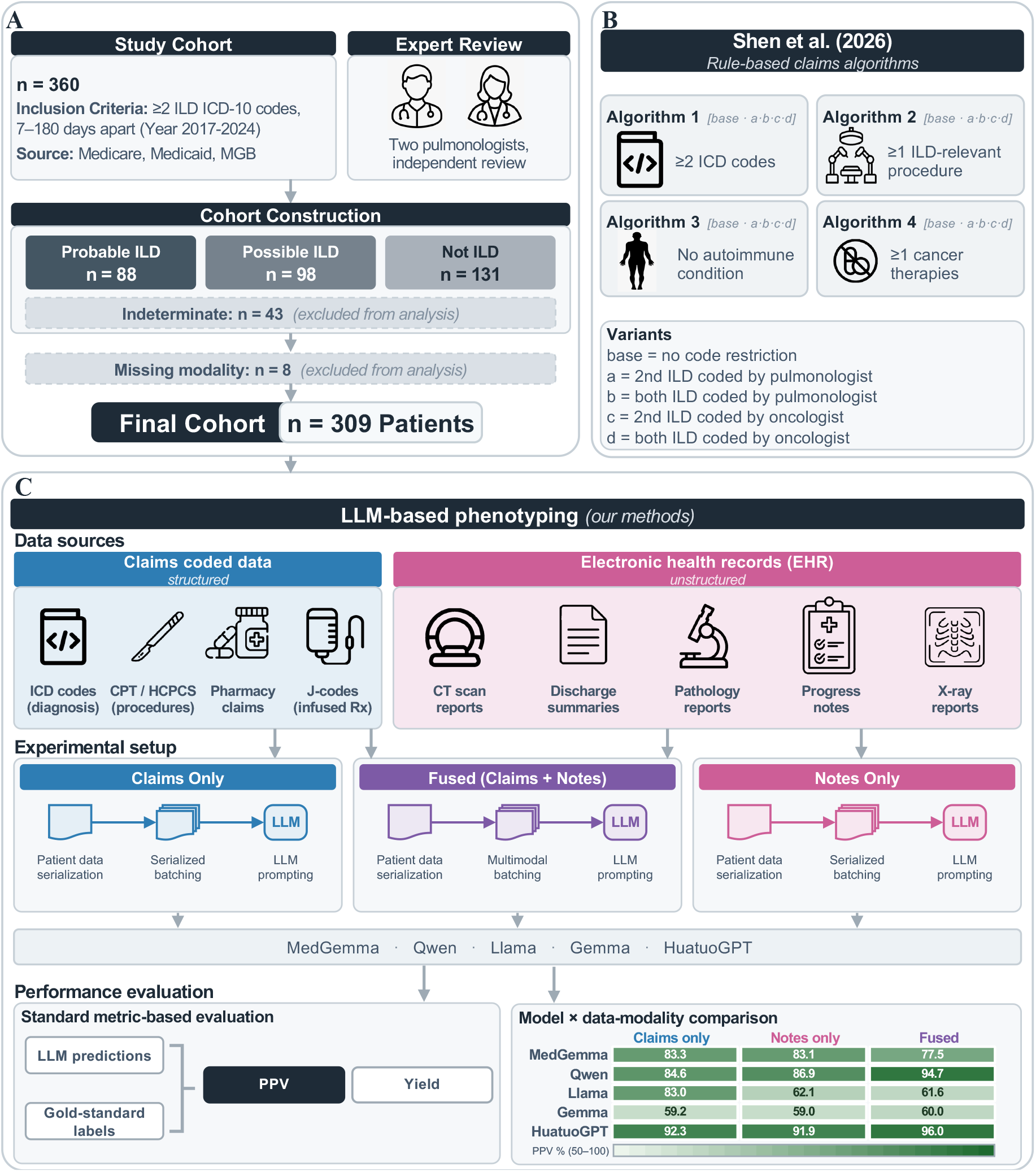
Study design. (**A**) The Mass General Brigham (MGB) oncology cohort of Shen et al. ^10^ (*n* = 360; entry criterion, at least two ILD diagnosis codes 7-180 days apart in patients with breast, lung, colorectal, or gynecologic cancer) was independently reviewed by two pulmonologists and adjudicated into Probable ILD (*n* = 88), Possible ILD (*n* = 98), Not ILD (*n* = 131), and Insufficient Evidence (*n* = 43; excluded). A further 8 patients without retrievable electronic health record data in the observation window were excluded, giving the final cohort of *n* = 309. (**B**) The 20 published claims-based algorithms of Shen et al. (Algorithms 1-4 and their pulmonologist- and oncologist-coded variants: a,b,c and d) are the rule-based comparator. (**C**) For each patient, linked claims and free-text notes within a 365 to +180-day window from the cohort entry date were collected, serialized and split into chunks for prompting. Twelve open-weight models spanning five families were evaluated against the reference standard using structured claims alone, clinical notes alone, and their fusion. All comparisons against the published algorithms were performed on identical patient sets using PPV and Yield as primary metrics.

For each patient, we extracted structured claims and free-text notes within a window of −365 to +180 days from CED, identical to the chart-review window used to generate the reference standard. We framed the task as retrospective phenotyping: given the records available to an expert reviewer, determine whether the patient had ILD at the index date.

### Data sources

We used structured claims from Medicare (January 1, 2016 to December 31, 2022), Medicaid (January 1, 2016 to December 31, 2021), and the MGB Health Plan (January 1, 2020 to December 31, 2024), comprising ICD-10-CM diagnosis codes, CPT and HCPCS procedure codes, and dispensed medications. Procedures included 42 Part-B-administered antineoplastic J-codes treated as ILD-relevant (Supplementary Table S2). Free-text data comprised typed clinical notes within the observation window. Note metadata were used to assign each note one of ten EHR note types: chest CT reports, pathology and biopsy reports, pulmonary function tests (PFTs), discharge summaries, chest X-rays (CXR), pulmonology encounters, progress notes, history-and-physical documents, ambulatory notes, and other documents.

Claim codes were serialized as text descriptions, each dated relative to the cohort entry date. Medications associated with drug-induced ILD were tagged inline with their risk level and drug class (for example, “[HIGH ILD risk: ICI]”), so that the model saw the risk alongside the drug rather than having to infer it (Supplementary Methods S1.2). Clinical notes were ordered by EHR note type, most informative for ILD first (chest CT, then pathology, then pulmonary function tests), so that these documents entered the first model input when a patient’s notes exceeded one context window (Supplementary Table S3). No disease-specific keyword filtering was applied. Patient demographics were included in every model input (Table 1). The study was approved by the MGB Institutional Review Board (Protocol #2017P002659), which waived the requirement for informed consent. The study protocol was not registered or published. Patients and the public were not involved in the design, conduct, or reporting of this study.

**Table 1.** Cohort characteristics (study cohort, *n* = 309). The Mass General Brigham oncology cohort of Shen et al. ^10^ restricted to patients with a definitive adjudicated label and at least one retrievable clinical note, enabling matched-cohort comparison across input modalities. Probable and Possible ILD are combined into the positive class for binary evaluation.

|  | Patients | % |
|---|---|---|
| study cohort | 309 | 100.0 |
| *Adjudicated label* |  |  |
| Probable ILD | 86 | 27.8 |
| Possible ILD | 97 | 31.4 |
| Not ILD | 126 | 40.8 |
| ILD-positive (Probable + Possible) | 183 | 59.2 |
| ILD-negative (Not ILD) | 126 | 40.8 |
| *Sex* |  |  |
| Female | 219 | 70.9 |
| Male | 90 | 29.1 |
| *Age, years* |  |  |
| Mean (SD) | 73.9 (8.4) |  |
| Median [IQR] | 73 [69-80] |  |
| Range | 36-94 |  |
| *Race* |  |  |
| White | 275 | 89.0 |
| Black or African American | 7 | 2.3 |
| Asian | 6 | 1.9 |
| Unknown | 21 | 6.8 |
| *Cancer type* |  |  |
| Lung | 136 | 44.0 |
| Breast | 99 | 32.0 |
| Colorectal | 44 | 14.2 |
| Ovarian | 22 | 7.1 |
| Endometrial | 8 | 2.6 |
| *Data substrate* |  |  |
| Medicare/Medicaid | 303 | 98.1 |
| MGB-HP | 6 | 1.9 |

### Patient-level aggregation through chunking

Because models tokenize the same text differently, each note was tokenized with the tokenizer of every evaluated model and assigned the largest of those token counts. Notes exceeding a single context window were then packed, in the fixed EHR note type priority order, into chunks of up to 123,964 tokens measured against that worst-case count, leaving the remainder of the 128,000-token window for the system prompt and the generated output. For fusion, the structured-claims block was included as a fixed prefix in every chunk. The notes-only and fused configurations contained the same clinical notes for each patient, although chunk boundaries could differ because the claims prefix occupied part of the available context.

The model generated one prediction per chunk. For binary evaluation, a patient was classified as positive if any chunk was labelled Probable or Possible ILD, following the human annotation rule. The three-class patient label was the highest-ranked chunk label, ordered as Probable ILD *>* Possible ILD *>* Not ILD. We compared this rule with three alternatives applied to the same chunk-level predictions (Supplementary Table S7): first chunk only, which uses the label of the first chunk, containing the highest-priority EHR note types; majority vote, which uses the most frequent label across a patient’s chunks; and maximum confidence, which uses the label of the chunk with the highest model-reported confidence. Per-patient token counts and chunk distributions are shown in Supplementary Fig. S2. We also examined how applying thresholds to model-reported ILD probabilities affected PPV and yield (Supplementary Fig. S3).

### Models and inference

We evaluated 12 diversed open-access LLMs spanning medical and general-purpose training, parameter scales, reasoning augmentation, and model families. These included MedGemma models with 4 and 27 billion parameters ^21^ and the general-purpose base model Gemma-3-27B. ^22^ We also evaluated Qwen3 models with 1.7, 4, 8, 14, and 32 billion parameters; ^23^ the reasoning-augmented HuatuoGPT-o1 medical models with 7 and 72 billion parameters; ^24^ and the general-domain non-thinkg models: Llama-3.3-70B-Instruct ^25^ and Qwen2.5-72B-Instruct models. ^26^ Details are presented in Supplementary Table S1.

Within each input configuration, all models received the same prompts and patient-data chunks. Prompts were developed on this cohort by the study team. Inference was zero-shot, without fine-tuning, retrieval, or in-context exemplars. We used greedy decoding (*T* = 0, with top-*p* and top-*k* sampling disabled), a fixed seed (42), no repetition penalty, a 128,000-token context, and tensor parallelism over four H100, served with vLLM (version 0.19.0).^27^ For each chunk, the model returned a structured JSON response containing a categorical label, an estimated probability that the patient had ILD, a self-rated confidence in that label, and a short rationale with supporting evidence. The configuration-specific system prompts are reproduced in Supplementary Table S8. All computation was performed on servers within the Mass General Brigham network.

### Evaluation

The primary measures were positive predictive value (PPV) and yield. PPV was the number of patients classified as ILD-positive according to the reference standard (i.e., manual chart review), divided by the number of patients classified as ILD-positive by the algorithm of interest. Yield was the number of patients classified as ILD-positive, divided by the number of patients evaluated (309). Like sensitivity, yield describes the breadth of case identification. We used yield because patients were pre-selected by ILD diagnosis codes, preventing assessment of overall detection sensitivity, including cases missed by this initial screening. We compared PPV–yield frontiers, defined by methods for which no alternative achieved equal or higher PPV and yield with a strict improvement in at least one measure.

To assess whether individual EHR note types supported improved ILD phenotyping, we evaluated chest CT reports, pulmonary function tests, discharge summaries, chest radiographs, and progress notes, each alone or combined with claims. MedGemma-27B was used throughout to hold the model constant while varying the EHR note type. For each EHR note type, the LLM and reference algorithms were evaluated on the same patients with that EHR note type, and LLM-to-rule ratios (*LLM_metric/rule_metric*) were calculated for PPV and yield. Detailed results are reported in Supplementary Tables S4–S6.

Each model also reported a probability that the patient had ILD. Because such probabilities could be used to rank patients for review or to set positivity thresholds, we assessed whether they were reliable. Discrimination was measured with the area under the receiver operating characteristic curve (AUROC), the probability that a randomly chosen patient with ILD receives a higher score than one without; 0.5 corresponds to chance, and values below 0.5 indicate that patients without ILD tend to receive higher scores. Calibration was measured with the expected calibration error (ECE), a measure commonly used for machine-learning classifiers:^28,29^ the average absolute difference between the reported probability and the observed proportion of patients with ILD at that probability, weighted by the number of patients; 0 indicates perfect calibration. We also examined PPV and yield across probability thresholds (Supplementary Fig. S3).

To assess implications for drug-safety studies, we projected how each classifier’s outcome mis-classification would attenuate a hypothetical drug–ILD association under non-differential mis-classification.^12,30^ Each classifier’s sensitivity and specificity, computed within the study cohort, were assumed to apply to the hypothetical study population and to be identical in exposed and unexposed patients. For a true odds ratio of 3.0 and ILD prevalence of 0.10 among unexposed patients, we reconstructed the expected misclassified two-by-two table and calculated the observed odds ratio, the sample size required to detect it with 80% power, and power across sample sizes for a cohort with equal numbers of exposed and unexposed patients, at *α* = 0.05. We also examined projected odds ratios across baseline prevalences of 0.05–0.20.

### Statistical analysis

Proportions are reported with Wilson 95% confidence intervals. Aggregation-strategy comparisons used 1,000 bootstrap resamples with a fixed seed (42). Paired differences in classification accuracy relative to Algorithm 4 (best non-specialist coded algorithm) were assessed using two-sided exact McNemar tests. For epidemiological projections, uncertainty in the within-cohort sensitivity and specificity was propagated using 2,000 Monte Carlo draws from Jeffreys-prior Beta posteriors to obtain 95% intervals.

Model inference used vLLM (version 0.19.0), ^27^ with tokenizers and chat templates loaded through Hugging Face Transformers. ^31^ Analyses were performed in Python 3.11 using pandas, NumPy, and SciPy. Reporting followed TRIPOD+AI and TRIPOD-LLM.^32,33^

## 3 Results

### Study cohort

The study cohort comprised 309 oncology patients with at least two claims-coded ILD diagnoses and retrievable clinical notes (Fig. 1; Table 1). After excluding 8 patients with missing modality, expert adjudication classified 86 patients as Probable ILD, 97 as Possible ILD, and 126 as Not ILD. Combining Probable and Possible ILD yielded 183 ILD-positive patients (59.2%).

### Zero-shot LLMs extend the PPV–yield frontier across input modalities

The LLMs extended the rule-based PPV–yield frontier under all three input configurations: claims alone, clinical notes alone, and their combination (fusion; Fig. 2A). Under claims alone, 10 of the 12 models were non-dominated, meaning that no published rule achieved both a higher PPV and a higher yield; the same held for 11 of 12 models under notes and 11 of 12 under fusion. The exceptions were models like Gemma-3-27B which classified a large fraction of the cohort as positive and therefore approached the cohort prevalence. This advantage was evident even without clinical notes. With claims alone, Qwen3-8B achieved 89.8% PPV at 15.9% yield, compared with 86.7% PPV at 4.9% yield for Algorithm 4a, the highest-PPV specialist-coded rule (Fig. 2B). The model therefore achieved a higher PPV while classifying more than three times the proportion of patients as positive, demonstrating that improved phenotyping was possible using claims alone.

**Figure 2.**
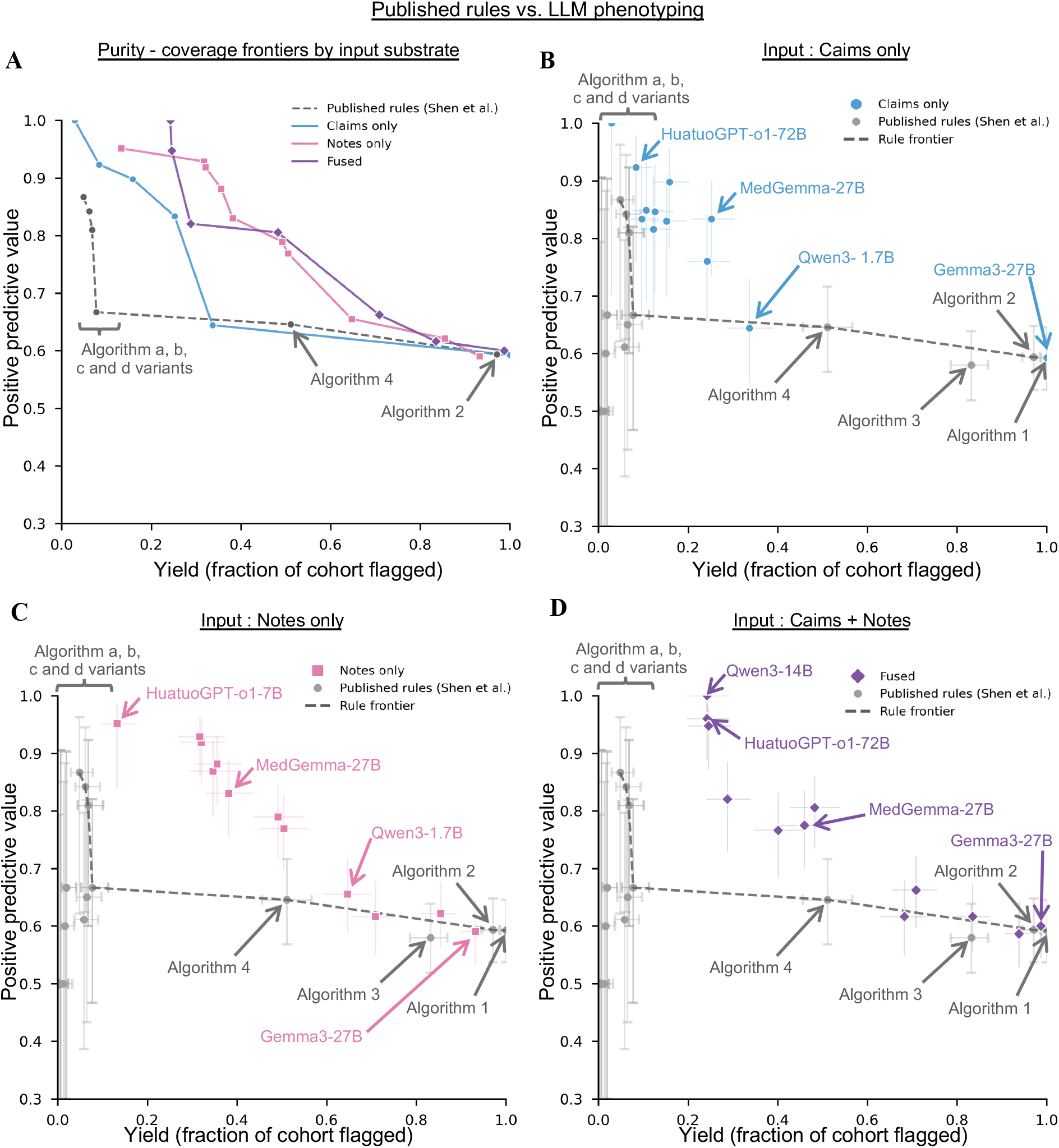
Zero-shot LLM phenotyping exceeds the rule-based PPV-yield frontier under every input data. PPV is plotted against yield, the fraction of the 309-patient cohort classified as ILD positive. In all panels, grey markers are the 20 published claims-based algorithms of Shen et al. ^10^ with Wilson 95% confidence interval crosses, and the grey dashed line traces their Pareto frontier, the set of rules for which no other rule achieves both higher PPV and higher yield. (**A**) The Pareto frontier of the 12 evaluated models under each input data, overlaid on the rule frontier; only non-dominated points are shown. All three model frontiers lie above the rule frontier across the range of achievable yields. (**B**-**D**) Individual models under each input data, with Wilson 95% confidence intervals: structured claims alone (**B**), clinical notes alone (**C**), and the two combined (**D**). Algorithm 1, which classifies every patient as positive, sits at the cohort prevalence (PPV 59.2% at 100% yield); the specialist-coded variants (a, b, c and d) cluster at high PPV and yields below 8%; Algorithm 4 is the best-performing rule with substantial yield (64.6% at 51.1%).

Clinical notes also supported high-PPV phenotyping at yields above those of the specialist-coded rules. Qwen3-14B achieved 92.9% PPV at 31.7% yield with notes alone (Fig. 2C). Adding claims increased its PPV to 100.0% (95% CI 95.1–100.0), while reducing yield to 24.3% (Fig. 2D). Other models offered higher-yield operating points. Qwen3-32B with fusion achieved 80.5% PPV and 48.2% yield, compared with 64.6% PPV and 51.1% yield for Algorithm 4, the best-performing rule. This represented a 15.9-percentage-point increase in PPV with a similar yield. Thus, the LLM advantage extended from highly selective case identification to phenotyping at yields close to the broader rule-based comparator.

### Document-restricted LLM phenotyping improves PPV over matched rule-based comparators

We next tested whether the advantage over rule-based phenotyping persisted when LLMs received a single EHR note type instead of the whole breadth of available notes. For that experiment, we fixed the LLM to be MedGemma-27B. Each comparison used the same patients with the relevant EHR note type. PPV point estimates exceeded those of Algorithm 4 in nine of ten evaluations across five EHR note types (Fig. 3A,B). Among patients with chest CT reports (*n* = 283), fusion achieved 80.5% PPV and 52.7% yield, compared with 64.8% PPV and 51.2% yield for Algorithm 4. Discharge summaries alone (*n* = 244) also achieved higher PPV than Algorithm 4, at 89.6% versus 63.7% (Fig. 3C,D). Because eligible populations differed across EHR note types, these findings do not establish which EHR note type performs best. The matched comparisons nevertheless show that the PPV advantage over rule-based phenotyping can persist when the model receives a single EHR note type.

**Figure 3.**
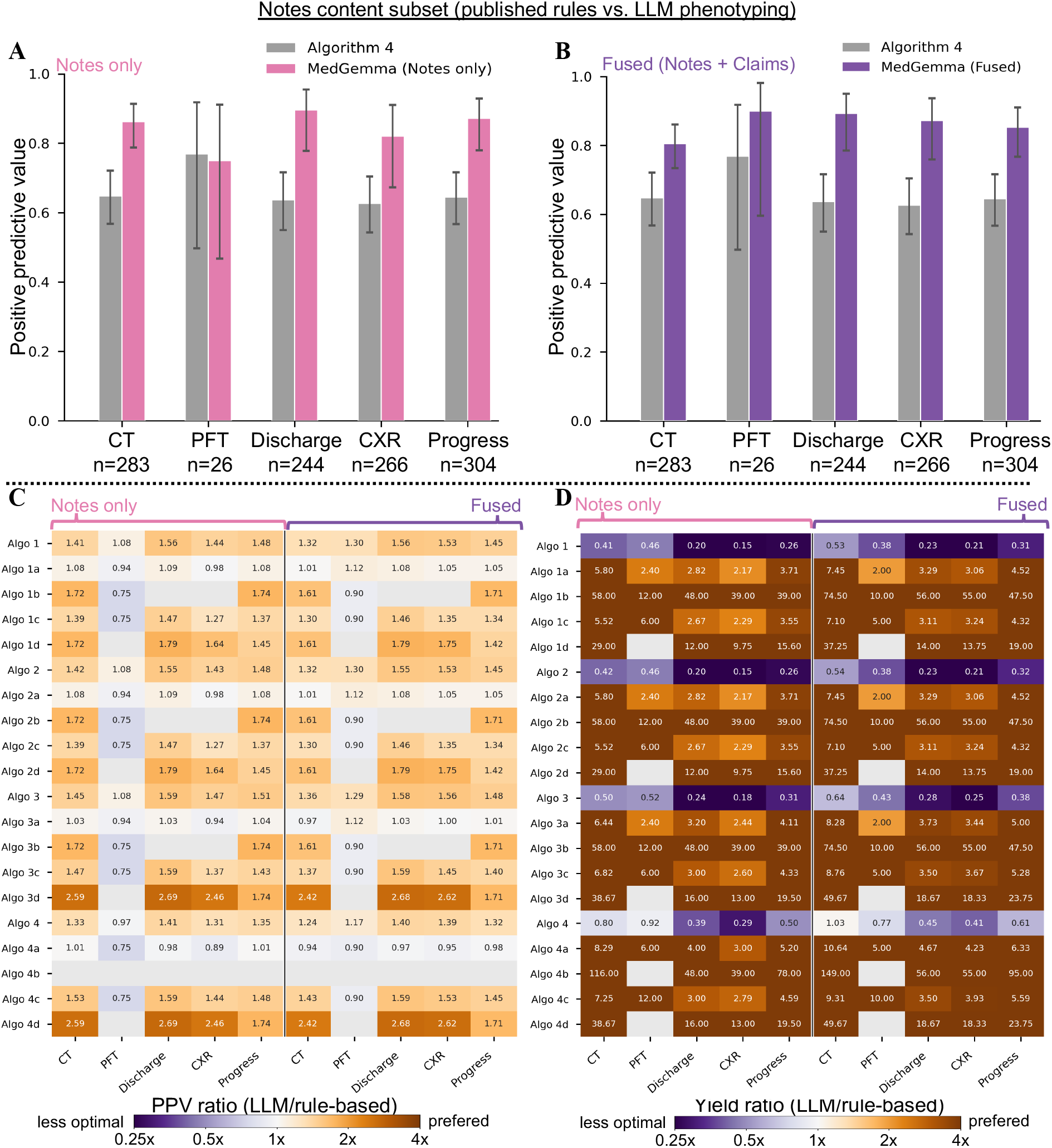
EHR note type ablation and matched-cohort head-to-head against every published algorithm. (**A**,**B**) PPV with Wilson 95% confidence intervals for the best non-specialist published rule (Algorithm 4, grey) versus the LLM under notes (**A**, pink) and Fused (**B**, purple), each restricted to the subcohort possessing the indicated EHR note type. Five EHR note types are shown: chest CT (*n* = 283), pulmonary function tests (*n* = 26), discharge summaries (*n* = 244), chest X-ray (*n* = 266), and progress notes (*n* = 304). The LLM exceeded the PPV of Algorithm 4 in nine of the ten document-by-configuration comparisons. **C**,**D**) Matched-cohort head-to-head ratios versus every published Shen rule. Rows, the 20 Shen algorithm variants; columns, the five EHR note types under each LLM configuration. Cell colour represents the ratio of LLM PPV (**C**) or yield (**D**) to the published rule; read jointly, the LLM dominates a rule where both ratios exceed 1. Ratios *>* 1 indicate the LLM outperforms. Cells where the metric cannot be computed (rule-based algorithm doesn’t flag any patient are masked grey).

### Clinical notes increase yield, while model choice shapes PPV

We next examined how input data and model choice shaped the PPV–yield trade-off. Replacing claims with clinical notes increased yield for 11 of 12 models (Fig. 4C; Supplementary Table S5). The exception, Gemma-3-27B, already classified every patient as positive with claims alone. Model choice also affected PPV. Under fusion, MedGemma-27B achieved 77.5% PPV, compared with 60.0% for its general-purpose base model, Gemma-3-27B (Fig. 4B). This higher PPV was accompanied by greater selectivity, with yields of 46.0% and 98.7%, respectively. Several general-purpose models also achieved higher PPV than MedGemma-27B, indicating that high PPV was not restricted to medically specialised models (Fig. 4A). Adding claims to notes further changed the operating point in different ways: some models gained PPV at lower yield, whereas others gained yield at lower PPV (Fig. 4D). Thus, clinical notes generally increased yield, while the resulting PPV–yield trade-off depended on the model and input combination. Performance stratified by cancer type is presented in Supplementary Fig. S4.

**Figure 4.**
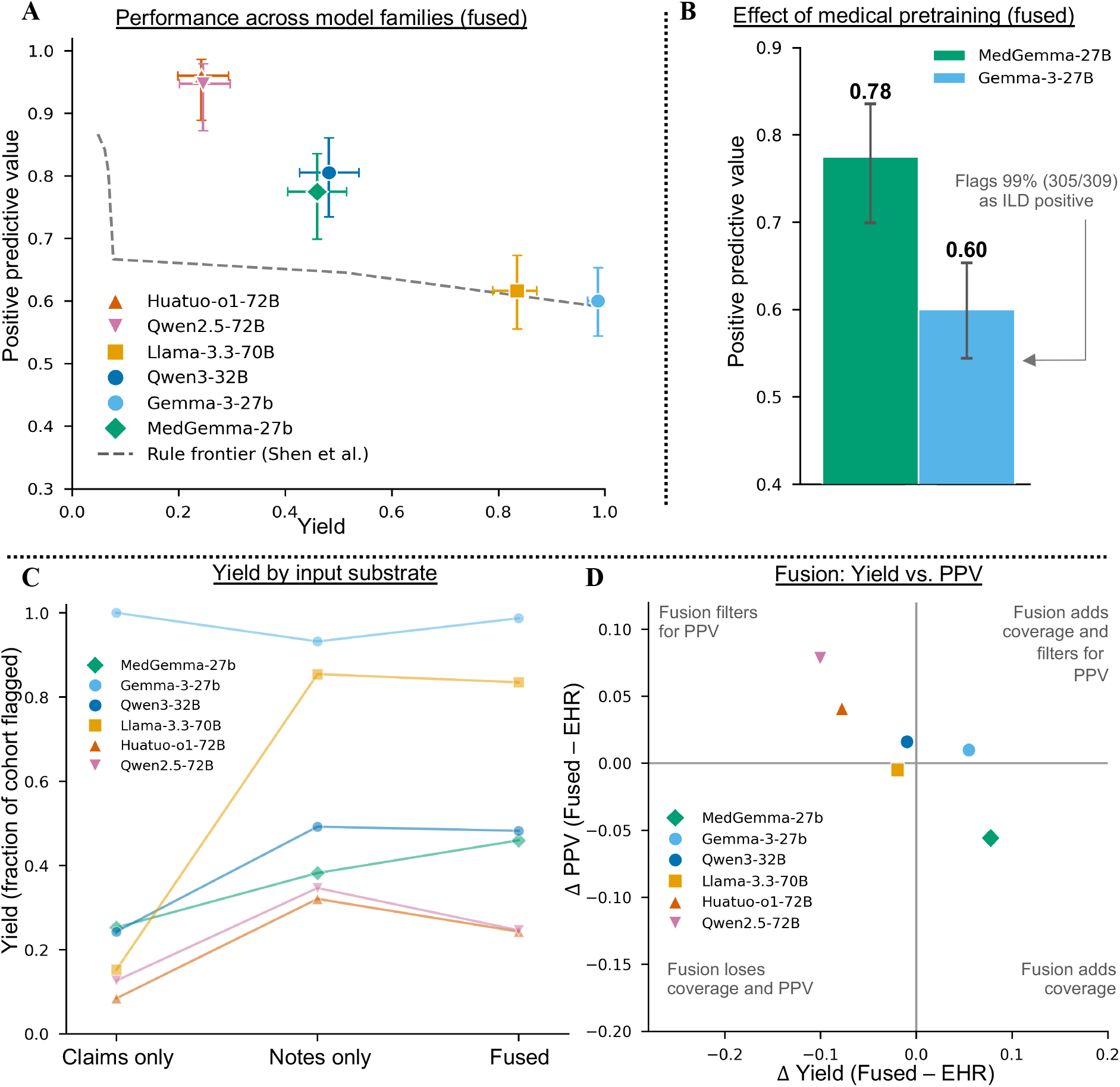
Determinants of LLM phenotyping performance across models and input configurations. (**A**) PPV versus yield for the largest model in each family under the Fused configuration on the 309-patient study cohort (MedGemma-27B, Gemma-3-27B, Llama-3.3-70B, HuatuoGPT-o1-72B, Qwen2.5-72B, Qwen3-32B); markers family-coloured per the legend, the non-dominated frontier of the published Shen rules ^10^ is overlaid (grey dashed line). The complete 12-model set and its per-cancer and scaling behaviour are given in Supplementary Fig. S6. (**B**) Matched medical-pretraining ablation: MedGemma-27B (green) versus its non-medical base Gemma-3-27B (sky-blue), which share architecture, parameter count and prompt, across PPV and yield with bootstrap 95% confidence intervals (*n* = 1,000, seed = 42). Gemma-3-27B is annotated “flags 99% (305/309)”: it labels virtually every patient ILD-positive, so its yield reaches 98.7% while its PPV stays at the cohort prevalence (60.0%). (**C**) Yield of every evaluated model under each input data (Claims only, Notes only, Fused); each line is one model, family-colored. Yield rises from claims to notes for every model except the degenerate Gemma-3-27B.(**D**) Change in yield versus change in PPV on moving from notes to fusion (Δ = Fused Notes), one point per model: upper-left, fusion acts as a PPV filter (yield falls, PPV rises); lower-right, fusion adds coverage. Adding structured claims to notes is therefore not uniformly coverage-maximizing but a re-reasoning step whose net effect depends on the model (per-patient decomposition in Supplementary Fig. S7 and Supplementary Table S9).

### Strong categorical performance does not ensure reliable probability estimates

We next examined whether the probabilities accompanying categorical predictions reliably reflected ILD status. Within each model-input evaluation, the emitted probabilities occupied only 3 to 13 distinct values, depending on the model and input, limiting the resolution of probability-based thresholds. Overall, high categorical PPV did not translate into reliable probability-based discrimination: Qwen3-14B achieved AUROCs of 0.300 with notes and 0.322 with fusion, below the 0.5 expected by chance, meaning that patients without ILD tended to receive higher probabilities than patients with ILD. Calibration was also poor. For MedGemma-27B under fusion, the expected calibration error was 0.238: reported probabilities differed from the observed proportion of patients with ILD by 24 percentage points on average (Supplementary Fig. S5). Thus, accurate identification of a high-PPV case set did not establish the reliability of the accompanying probabilities, which require separate validation before use as estimates of uncertainty.

### LLM phenotyping is projected to reduce effect attenuation and sample-size requirements

We finally assessed whether improved phenotyping could preserve drug–ILD associations and reduce the sample size required to detect them. We modeled a cohort with equal numbers of exposed and unexposed patients, with a hypothesised true odds ratio (OR) of 3.0 and baseline ILD prevalence of 0.10. Each classifier’s measured sensitivity and specificity were used to project the observed OR under non-differential outcome misclassification (Methods). The best rule-based algorithm (Algorithm 4) reduced the observed OR to 1.07, nearly erasing the true association, whereas Qwen3-14B preserved it to increasing degrees: 1.31 with claims, 1.80 with notes, and 2.67 with fusion (Fig. 5C). LLM with fusion input therefore preserved the largest proportion of the hypothesised association. The lower projected attenuation relative to Algorithm 4 persisted across the examined baseline prevalences of 0.05–0.20.

**Figure 5.**
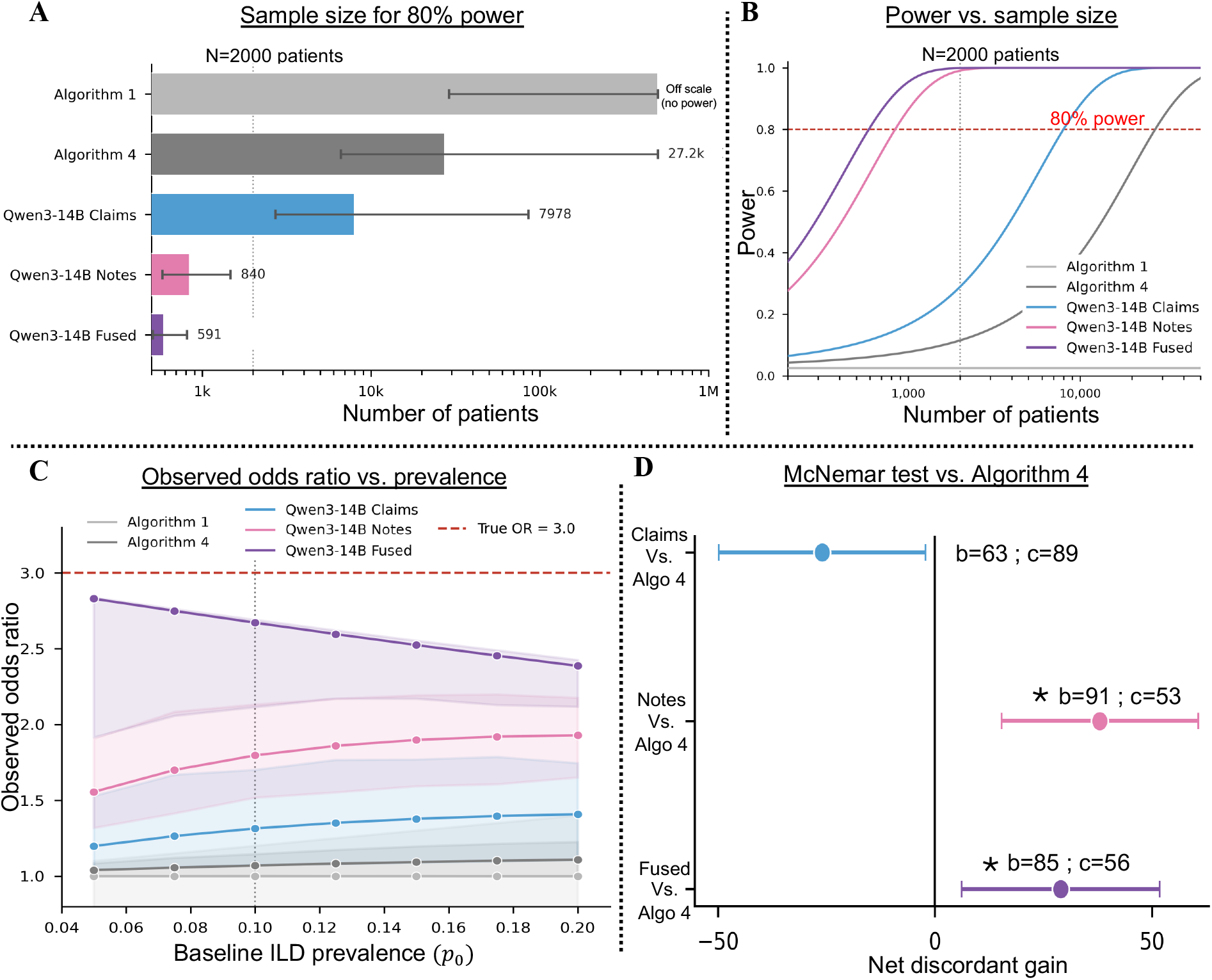
Statistical power and matched-cohort efficiency of the LLM phenotype. For a hypothesised drug-ILD association with true odds ratio = 3.0, baseline ILD prevalence *p*_0_ = 0.10, and a cohort with equal numbers of exposed and unexposed patients, each classifier’s measured operating characteristics were propagated through the Copeland ^12^ non-differential outcome-misclassification formula to obtain the expected observed odds ratio, required sample size for 80% power, and power-sample-size curve. (**A**) Total sample size *N* (exposed + unexposed, 1:1) required to detect the effect at 80% power, by classifier. Error bars are Monte-Carlo 95% confidence intervals derived from Jeffreys-prior Beta posteriors on the misclassification-model inputs (*n* = 2,000 samples). Algorithm 1 sits off the plotted scale (degenerate: flags every patient, observed OR = 1). (**B**) Power-versus-*N* curves for each classifier, with reference lines at *N* = 2,000 (typical observational cohort) and at 80% power. At *N* = 2,000, Qwen3-14B Fused reaches 99.9% power, Qwen3-14B Notes 99.1%, Qwen3-14B Claims 28.9%, Algorithm 4 11.5%, and Algorithm 1 2.5%. (**C**) Robustness of the observed odds ratio to the assumed baseline ILD prevalence (*p*_0_ ∈ [0.05, 0.20]) at true OR = 3.0; shaded bands, Monte-Carlo 95% confidence intervals. (**D**) Paired head-to-head against the best published rule (Algorithm 4) on the 309-patient study cohort. For each LLM configuration, the McNemar discordant counts (*b*, LLM correct and rule 95% confidence interval 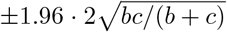 and the two-sided exact McNemar *p*-value. Vertical line at zero (no difference). Qwen3-14B Notes and Qwen3-14B Fused significantly outperform Algorithm 4 (*p* = 0.0019 and *p* = 0.018); Qwen3-14B Claims performs significantly worse (*p* = 0.042), reflecting its low yield: at 9.7% it identifies far fewer true cases than Algorithm 4 despite its higher PPV.

These differences translated into substantially lower projected sample-size requirements. At 80% power (*α* = 0.05), Algorithm 4 required approximately 27,200 patients. The corresponding requirements for Qwen3-14B were 7,978 with claims, 840 with notes, and only 591 recruted patients with fusion (Fig. 5A,B). The claims-only configuration therefore reduced the required sample size even without linked clinical notes, while fusion reduced it from tens of thousands of patients to hundreds. Under the specified assumptions, these phenotypes preserved more of the exposure-outcome association and enabled its detection in substantially smaller studies.

## 4 Discussion

In this matched evaluation of 309 oncology patients, selected LLMs identified ILD cases with higher PPV than the reference algorithms with similar or greater yields. The advantage was observed with the input of claims alone, clinical notes alone, and the fusion. Previous studies have shown that zero-shot LLMs can outperform rule-based methods for phenotyping chronic conditions,^19^ and that supervised machine-learning classifiers can outperform code-based rules for ILD identification. ^6^ Our study extends this work by comparing models and rules on the same expert-adjudicated patients and examining the implications for association studies. The principal advance is an improvement in the PPV–yield trade-off compared with 20 rule-based claim algorithms, ^10^ with potential consequences of reducing number of cases identified or a reduced sensitivity.

The projected epidemiological benefit gives this classification improvement a practical research consequence. Outcome misclassification can attenuate associations and increase the sample size needed to detect them. ^12,13^ Under the modelled scenario, the best LLM configurations required far fewer patients than the best rule-based algorithm to detect the same association. Improved automated detection of a phenotype could therefore make associations detectable in cohorts of a size that rule-based ascertainment would leave underpowered. The result depends on the classifier’s sensitivity and specificity and on the assumed prevalence; PPV alone does not determine the projected benefit.

The model and input comparisons clarify how such gains might be obtained. Clinical notes generally increased yield, while model choice altered the balance between PPV and the proportion classified positive. MedGemma’s higher PPV than its general-purpose base model was accompanied by greater selectivity, suggesting that medical pretraining shifted the operating point rather than uniformly improving classification. General-purpose models also achieved high PPV, and larger models generally performed better. These findings support evaluating candidate models empirically for the intended phenotype. Fusion also changed which cases were identified, gaining some detections and losing others relative to the separate inputs. Several individual EHR note types supported higher PPV than rules evaluated in the same eligible patients. Useful evidence could therefore be recovered from a single EHR note type.

The claims-only result is particularly relevant to administrative databases without linked clinical notes. Using open-weight models, we performed all inference within institutional infrastructure, providing a practical basis for analyzing protected records. Zero-shot inference avoided task-specific model training, although pipeline development and validation still required clinical expertise. The operating point could also be adjusted after inference by changing how a patient’s chunk-level predictions were combined, which could support different requirements for case confirmation or cohort assembly. The categorical label, rather than the emitted probability, should be the basis for case definition: using LLM outputs to rank patients for human review would require uncertainty estimates that these models did not provide.

This study has several limitations. First, this was a single-institution study of oncology patients already flagged by ILD codes. The findings therefore concern retrospective ILD ascertainment and do not independently validate prospective prediction or attribution to a particular drug. Performance in other health systems, and in larger populations not pre-selected by ILD codes, remains unknown. Second, high PPV often accompanied selective classification. We do not claim the PPV-yield phenotyping tradeoff solved, but merely found a way to improve its limitations across several patient input data. Third, we compared LLMs only with already deployed rule-based algorithms. Supervised machine-learning classifiers trained on claims or notes were not evaluated and could match or exceed zero-shot LLMs where labelled training data are available. Finally, the projections assume that sensitivity and specificity estimated in this enriched cohort apply to populations with lower baseline prevalence, and that misclassification is non-differential by drug exposure. Because model inputs included drug exposures annotated with their ILD risk, classification errors could differ by exposure. These assumptions were not tested in an actual association study. External validation and empirical drug–outcome analyses are needed to determine whether the projected gains are realised.

In summary, our findings suggest that LLMs achieved more favorable trade-offs between case identification and diagnostic precision than published claims-based algorithms, across different data input settings: claims only, EHR note only, or claims-EHR-note combined. Importantly, these improvements were achievable even with claims alone, without requiring linked clinical notes. Under specified outcome-misclassification assumptions, improved ascertainment was projected to reduce effect attenuation and substantially lower sample-size requirements, potentially enabling drug-safety studies that would otherwise be underpowered. Further advances through few-shot learning, ^34^ knowledge-enhanced inference, ^35,36^ and retrieval-augmented generation may improve phenotyping performance beyond zero-shot approaches and warrant future investigation.^37^ More broadly, our findings highlight the potential of LLM-based phenotyping to address outcome misclassification as a fundamental limitation of real-world evidence generation, enabling more reliable and efficient drug-safety and comparative effectiveness research.

## Contributors

GL, SS, and JY conceived the study. GL performed the analyses and wrote the manuscript. BG and PA contributed data, clinical adjudication, and methodological input. KJL, SS and JY are senior authors and supervised this study. GL and BG accessed and verified the underlying data. All authors had full access to the data in the study and accepted responsibility to submit for publication. All authors revised the manuscript and approved the final version.

## Supporting information

Completed TRIPOD+AI reporting checklist

Completed TRIPOD-LLM reporting checklist

## Data Availability

Patient-level data cannot be shared because of HIPAA regulations and Mass General Brigham Institutional Review Board restrictions. Aggregate results are reported in the manuscript and supplementary appendix. All analysis code is available at https://github.com/YLab-Open/ild-llm-phenotyping.

## Data sharing

Patient-level data cannot be shared because of HIPAA regulations and Mass General Brigham IRB restrictions. Aggregate results and all analysis code are available in the code repository.

## Code availability

The source code for the full analysis is available at https://github.com/YLab-Open/ild-llm-phenotyping under an MIT license.

## Acknowledgements

We thank the Shen et al. team for providing the patient-level algorithm-flag table, and the clinical reviewers and data-engineering staff who supported cohort construction. ChatGPT (OpenAI) was used solely for grammar checking and language refinement. All AI-assisted edits were reviewed and approved by the authors, who take full responsibility for the content of the manuscript.

