## Supplementary Methods, Figures S1-S7 and Tables S1-S9 for "Identifying interstitial lung disease with large language models on clinical notes and claims"

##### S1.1 Observation window, input configurations, and deterministic prompt assembly

For every patient we assembled, separately for each input configuration, a single deterministic text prompt from the structured claims and free-text notes falling within the −365 to +180 day observation window around the cohort entry date (CED), the same window used by the chart reviewers who generated the reference standard. Three configurations were evaluated on the identical 309-patient cohort: structured claims only; clinical notes (EHR) only; and Fused, claims and notes jointly. The three share a common serialization stack, so that a given patient's demographics and claims are rendered identically wherever they appear. The Claims configuration always fit in a single chunk; the Notes and Fused configurations were splits into chunks as required.

Each prompt opens with a fixed demographics header giving the index date (CED) followed by the patient's age at index (integer years), sex, race, insurance, and cancer type. The header is followed by the structured-claims block (Supplementary Methods S1.2) and, where applicable, the clinical-notes block (Supplementary Methods S1.3). The same patient data were substituted into the configuration-specific system prompts reproduced verbatim in Supplementary Table S8; no exemplars, retrieval, or task-specific tuning were used.

Token budgets were defined against the 128,000-token context shared by all models. From this we reserved 1,536 tokens for the generated answer, 1,500 tokens for the system prompt and chat-template overhead, and a 1,000-token safety buffer, leaving 123,964 tokens of patient content per chunk. Because models tokenize text differently, every note and fixed prefix was tokenized with the tokenizer of every evaluated model, and the largest count was used. Per-patient prompt-token and chunk-count distributions for the three configurations are shown in Supplementary Fig. S2.

##### S1.2 Structured-claims serialization

The structured-claims block renders a patient's administrative record as time-anchored clinical text. Every event is expressed relative to the CED and placed in one of three sections, recorded prior to, on, or after the index date, with each event's day offset written as "day $-N$", "day 0", or "day $+N$". Offsets are ordered most-recent-first before the index date and earliest-first after it. The block is built in four parts.

*Index ILD codes.* The block opens with the one or two ICD-10 ILD diagnosis codes that triggered cohort entry, each shown with a curated description for the canonical ILD codes (for example J84.9, J84.89, J70.4) and the institutional ICD description otherwise.

*Diagnoses.* All ICD-10 diagnoses in the window are listed with their descriptions and section placement.

*Procedures.* Procedure codes are split into two streams. ILD-relevant diagnostic procedures are retained in full temporal detail: chest CT, chest radiography, pulmonary function testing, bronchoscopy and endobronchial or transbronchial biopsy, CT-guided and open lung biopsy, bronchoalveolar lavage and thoracentesis, the six-minute walk test, echocardiography and right-heart catheterization (relevant to pulmonary hypertension), positron-emission tomography, and the Part-B administered antineoplastic J-codes detailed below. Procedure labels are location-independent, so the same procedure code recorded at different clinic sites is represented once rather than counted as new information. The second stream, routine laboratory monitoring, comprises codes in the 800xx to 899xx pathology and laboratory CPT range, the Q99x HCPCS range, and the institutional "EPRC:" laboratory prefix (excluding any code on the ILD-relevant list); to keep high-volume routine testing from displacing diagnostic signal, these are compacted into a summary giving the number of unique laboratory tests, the total order volume, the temporal span, and up to thirty test names with per-test order counts.

*Medications and ILD-risk flagging.* Dispensed medications are listed by name and section. Each patient's ILD-risk exposures, drawn from a precomputed drug-exposure table that records a risk level and drug class for every matched agent, are flagged inline (for example, "[HIGH ILD risk: ICI]"), so the model sees the risk annotation in context rather than having to infer it. Three summary counts (ILD-relevant procedures, routine laboratory tests, and ILD-risk medications) are recorded per patient as serialization metadata.

The Part-B administered antineoplastic J-codes that the serializer treats as ILD-relevant procedures, retained in full rather than compacted with routine procedures, are the fixed set listed in Supplementary Table S2, grouped by drug class.

##### S1.3 Clinical-note tiering and chunking

*Note selection and tiering.* All typed clinical notes in the MGB EHR within the observation window were eligible; no disease-specific keyword filtering was applied, so notes were selected by EHR note type alone. Each note was assigned one of ten EHR note types from its metadata. For serialization these types are assigned a packing priority that reflects diagnostic yield for ILD, so that higher-yield documents are placed first when context is limited (Supplementary Table S3).

*Per-note preparation.* Each note was anchored to the CED to obtain a relative day; notes with unparseable dates or empty text were dropped. Note text was tokenized with every evaluated model's tokenizer, and each note was assigned the largest count.

*Chunking.* Because the full note set for many patients exceeds a single context, notes were packed into chunks by a greedy algorithm. Notes were ordered by packing rank (highest-yield class first) and, within a class, by relative day (most recent first), then added to the current chunk until the next note would exceed the per-chunk budget, at which point a new chunk was opened. The per-chunk budget was the global content budget (123,964 tokens; Supplementary Methods S1.1) minus the tokens consumed by the fixed material re-shown in every chunk: in the Notes configuration, the demographics header (with a 200 token allowance for section headers); in the Fused configuration, the demographics header plus the complete structured-claims block, re-presented as a fixed prefix in every chunk. Consequently, for a given patient the set of notes shown is identical between the Notes and Fused configurations; only their partitioning into chunks differs, because in the Fused configuration the claims prefix consumes part of each chunk's budget. This preserves the matched comparison between the two note-bearing configurations.

*Within-chunk display order.* Although notes are packed in priority order, they are rendered for the model in chronological order: within each chunk the notes are regrouped by timing into sections recorded prior to, on, and after the index date and sorted by relative day, each under a header giving its note type and signed day offset. Packing order therefore governs only which notes share a chunk, not the order in which the model reads them.

*Patient-level aggregation.* The model produced one prediction per chunk; these were combined into a single patient-level label by the positivity-maximising rule described in the main-text Methods, with the alternative aggregation rules and their operating points characterised in Supplementary Table S7.

#### Supplementary Figures

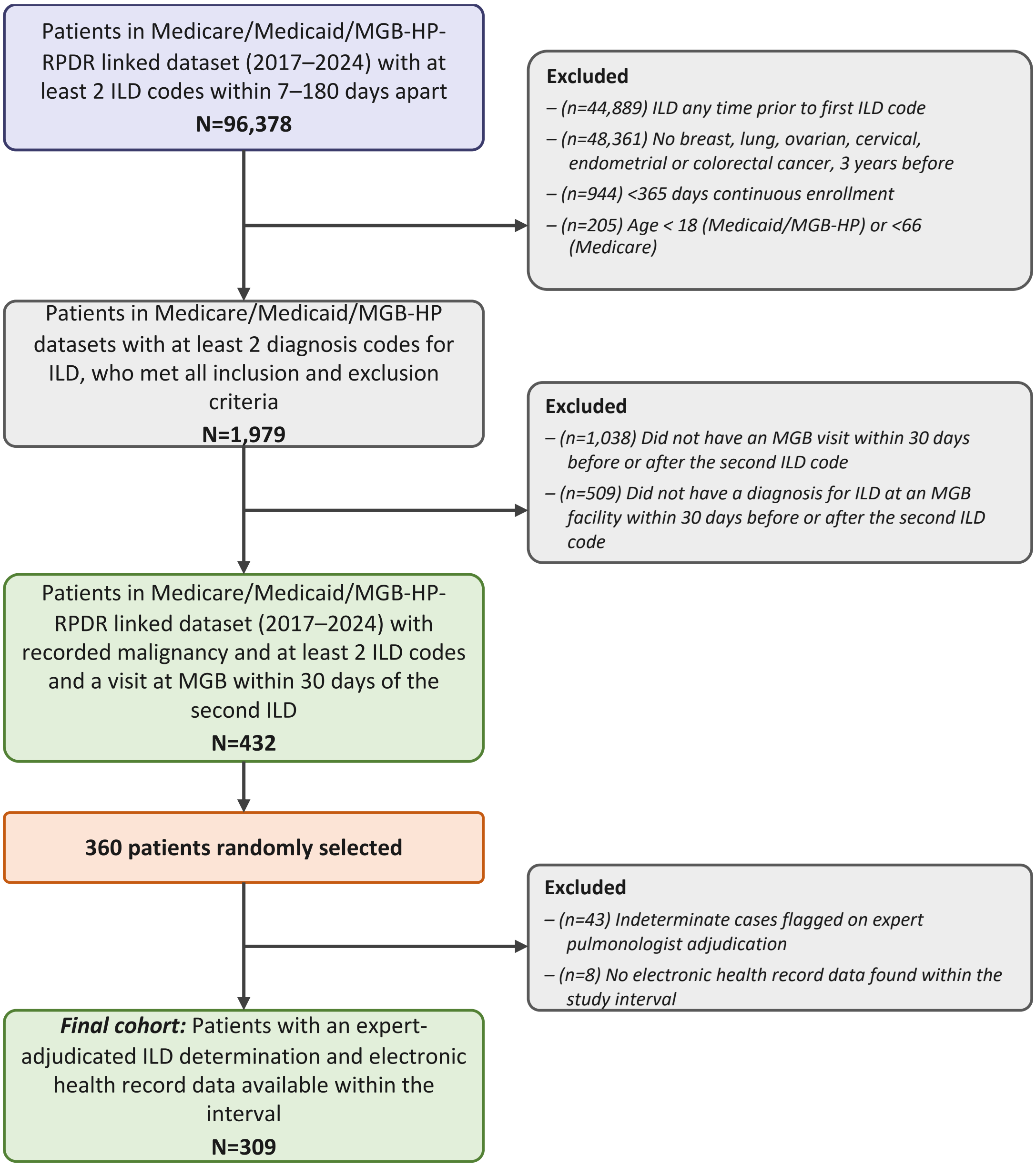


**Figure S1. Cohort assembly flow diagram.** Construction of the 309-patient study cohort used for all experiments and evaluation.

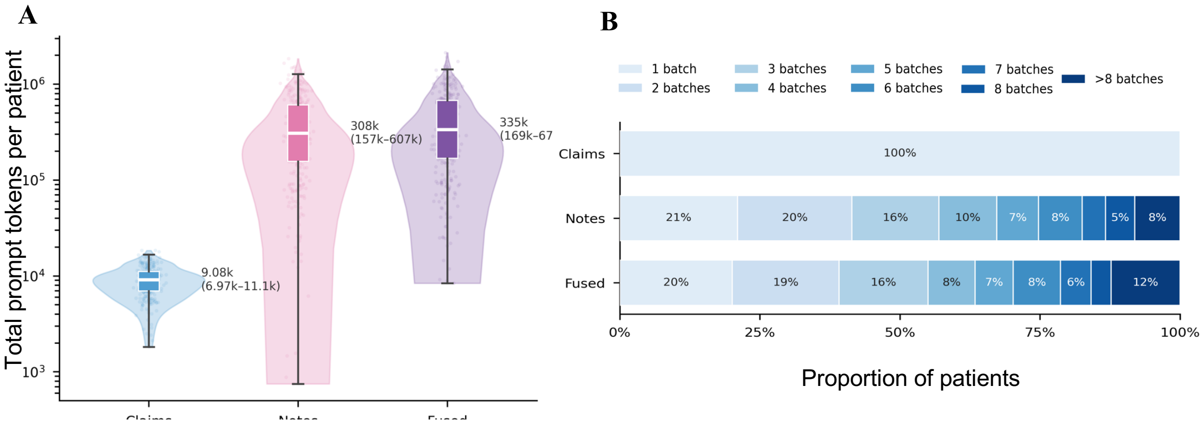


**Figure S2. Per-patient prompt token footprint and chunk distribution across the three configurations.** (**A**) Distribution of total prompt tokens consumed per patient under the Claims (blue, $n = 309$), Notes (pink, $n = 309$), and Fused (purple, $n = 309$) configurations. Annotation alongside each violin reports the median and IQR. (**B**) Distribution of $n_{\text{chunks}}$ per patient as a stacked-proportion bar (bins 1 through 8, plus >8), illustrating how often patients required more than one chunk under each configuration. All Claims-configuration patients fit in a single chunk; Notes and Fused distribute mass across larger chunk counts as the fixed claims prefix repeats per chunk in Fused.

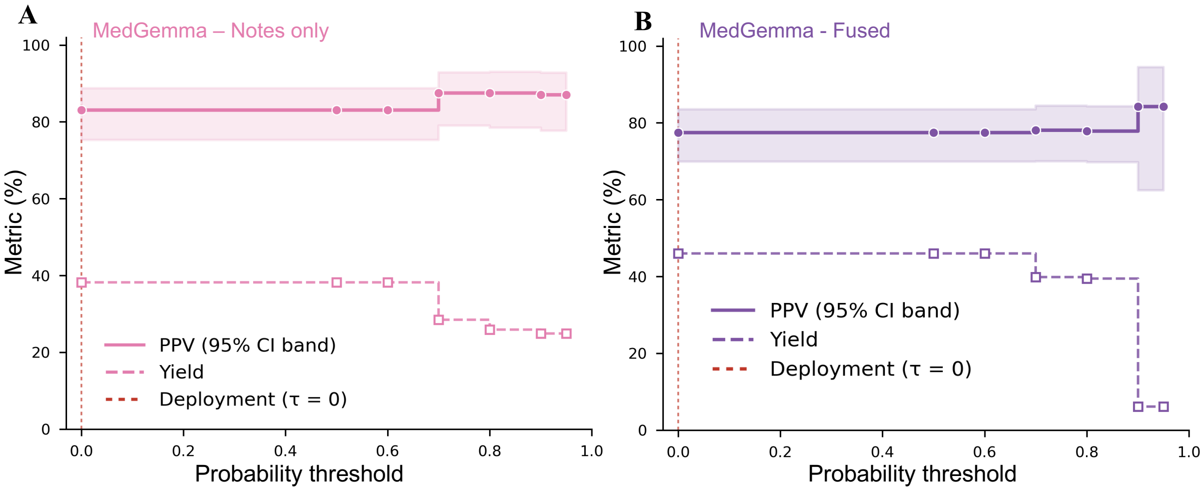


**Figure S3. Probability-threshold operating curves on the 309-patient study cohort.** Step plots of PPV (solid line, 95% bootstrap confidence interval as shaded band) and yield (dashed line) as the per-chunk positivity threshold $\tau$ is swept from 0 to 0.95 under the Notes (**A**; pink) and Fused (**B**; purple) configurations. The step shape reflects the discrete probability output of the model, which emits values in a small quantized set. The deployment threshold $\tau = 0$ (plain OR aggregation) is marked by the red dashed vertical line. Tightening the threshold to $\tau = 0.7$ gains only +4.4 pp PPV under Notes and +0.6 pp under Fused, at a cost of −9.7 pp and −6.1 pp yield respectively. Under Fused, PPV rises further to 84.2% at $\tau = 0.9$, but only by discarding an additional fifth of the classified cases. Thresholding the emitted probability therefore buys little purity for the coverage it removes, consistent with the poor discrimination of those probabilities reported in the Results, and $\tau = 0$ was used for all analyses.

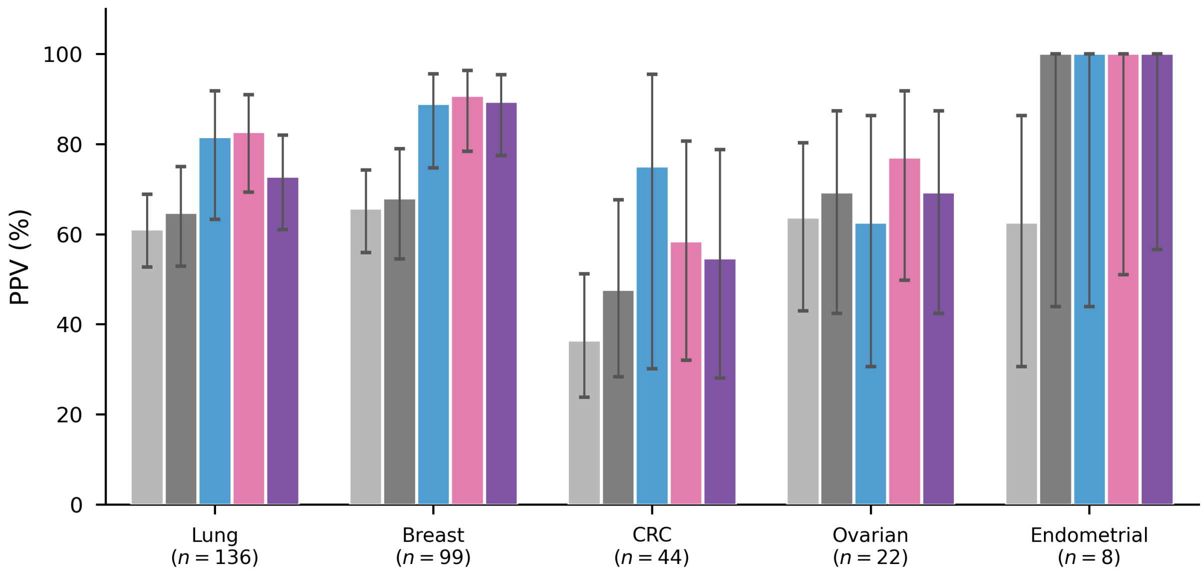


**Figure S4. Per-cancer-cohort head-to-head comparison.** Positive predictive value (PPV) of the three LLM configurations (Claims, Notes, Fused) and the two principal reference rules (Algorithm 1 and Algorithm 4), stratified by cancer subgroup on the 309-patient study cohort (lung, breast, colorectal, ovarian, endometrial). Error bars, Wilson 95% confidence intervals.

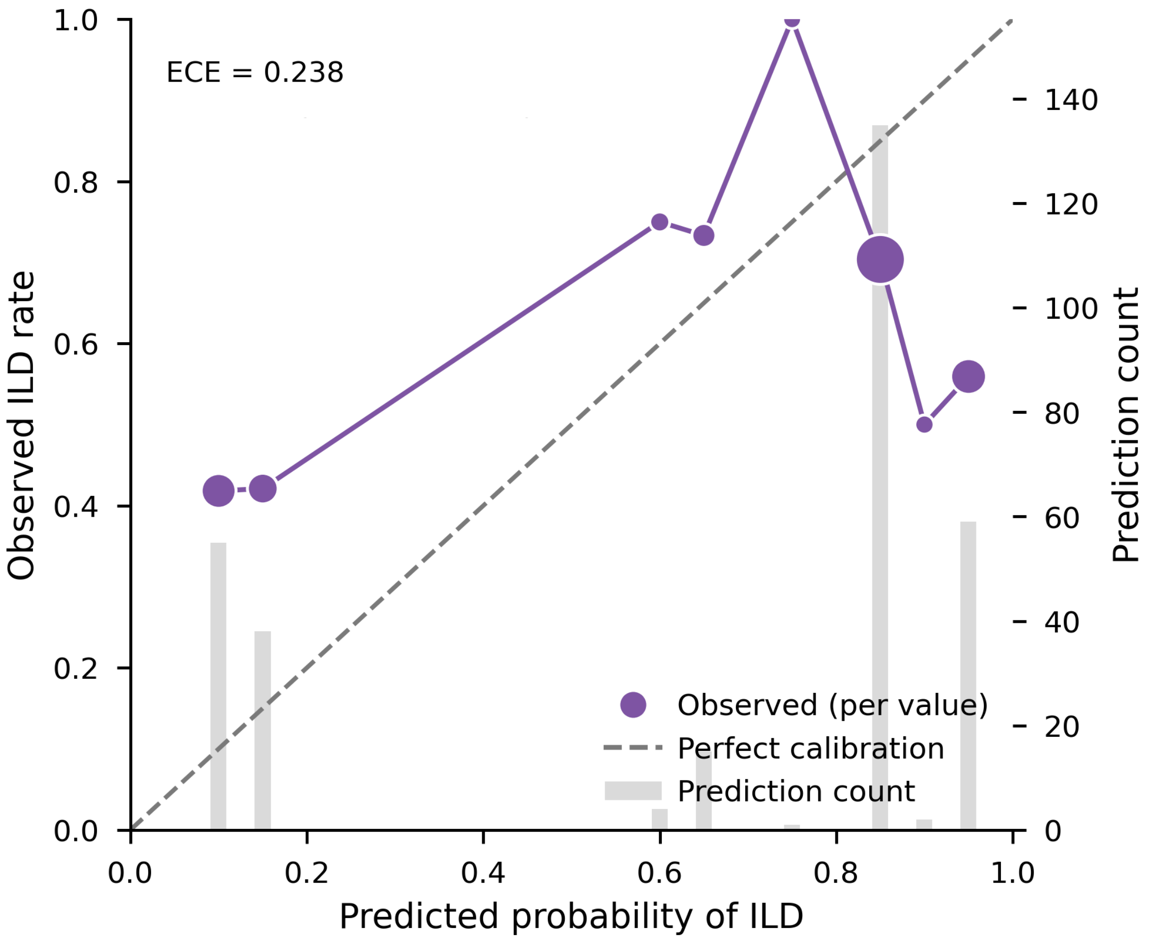


**Figure S5. Reliability diagram for MedGemma-27B under the Fused configuration on the 309-patient study cohort.** Each patient's ILD probability is the value emitted with the chunk that determined the patient's label. Purple points give, for each distinct probability value the model emitted, the fraction of patients at that value who were reference-positive (left axis). The number of patients at each value is shown both by marker area and, on the right axis, by the grey bars. The dashed diagonal denotes perfect calibration. The model emits a small set of discrete probability values rather than a continuous distribution (expected calibration error, 0.238).

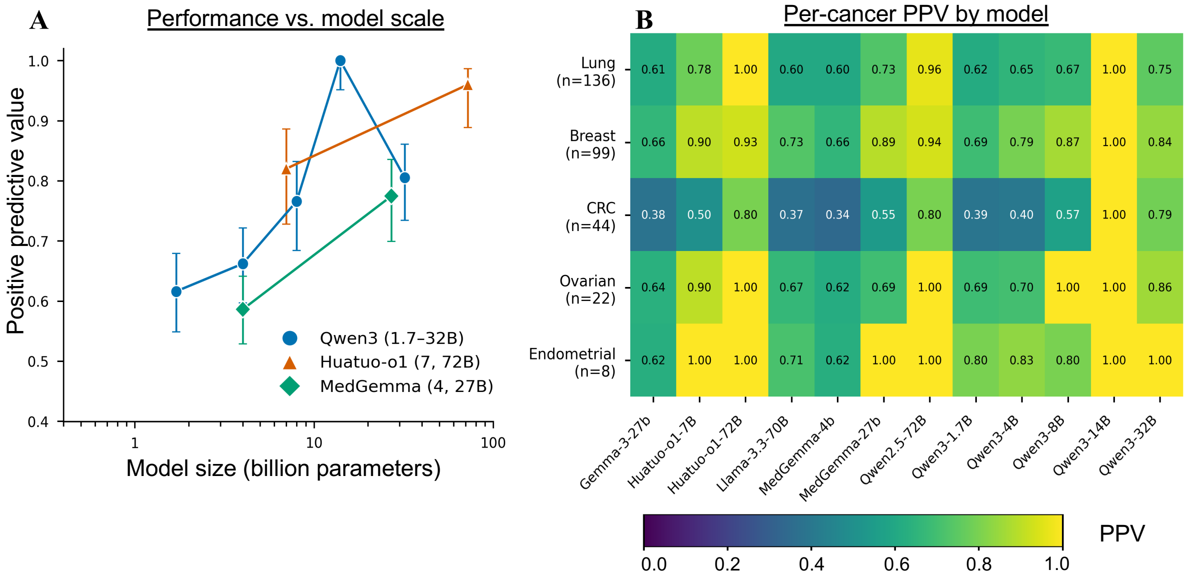


| | Gemma-3-27b | Huatuo-o1-7B | Huatuo-o1-72B | Llama-3.3-70B | MedGemma-4b | MedGemma-27b | Qwen2.5-72B | Qwen3-1.7B | Qwen3-4B | Qwen3-8B | Qwen3-14B | Qwen3-32B |
|---|---|---|---|---|---|---|---|---|---|---|---|---|
| Lung (n=136) | 0.61 | 0.78 | 1.00 | 0.60 | 0.60 | 0.73 | 0.96 | 0.62 | 0.65 | 0.67 | 1.00 | 0.75 |
| Breast (n=99) | 0.66 | 0.90 | 0.93 | 0.73 | 0.66 | 0.89 | 0.94 | 0.69 | 0.79 | 0.87 | 1.00 | 0.84 |
| CRC (n=44) | 0.38 | 0.50 | 0.80 | 0.37 | 0.34 | 0.55 | 0.80 | 0.39 | 0.40 | 0.57 | 1.00 | 0.79 |
| Ovarian (n=22) | 0.64 | 0.90 | 1.00 | 0.67 | 0.62 | 0.69 | 1.00 | 0.69 | 0.70 | 1.00 | 1.00 | 0.86 |
| Endometrial (n=8) | 0.62 | 1.00 | 1.00 | 0.71 | 0.62 | 1.00 | 1.00 | 0.80 | 0.83 | 0.80 | 1.00 | 1.00 |

**Figure S6. Additional model-comparison analyses (Fused configuration, 309-patient study cohort).** (**A**) Within-family scaling of PPV against parameter count on a log axis, for the Qwen3 family (1.7-32B), the HuatuoGPT-o1 medical family (7B, 72B), and the MedGemma family (4B, 27B); PPV with bootstrap 95% confidence intervals ($n$ = 1,000, seed = 42). PPV generally increased with parameter count within each family. (**B**) Per-cancer PPV heatmap. Rows are cancer subgroup (lung, breast, colorectal, ovarian, endometrial); columns are all the evaluated open-weight models grouped by family.

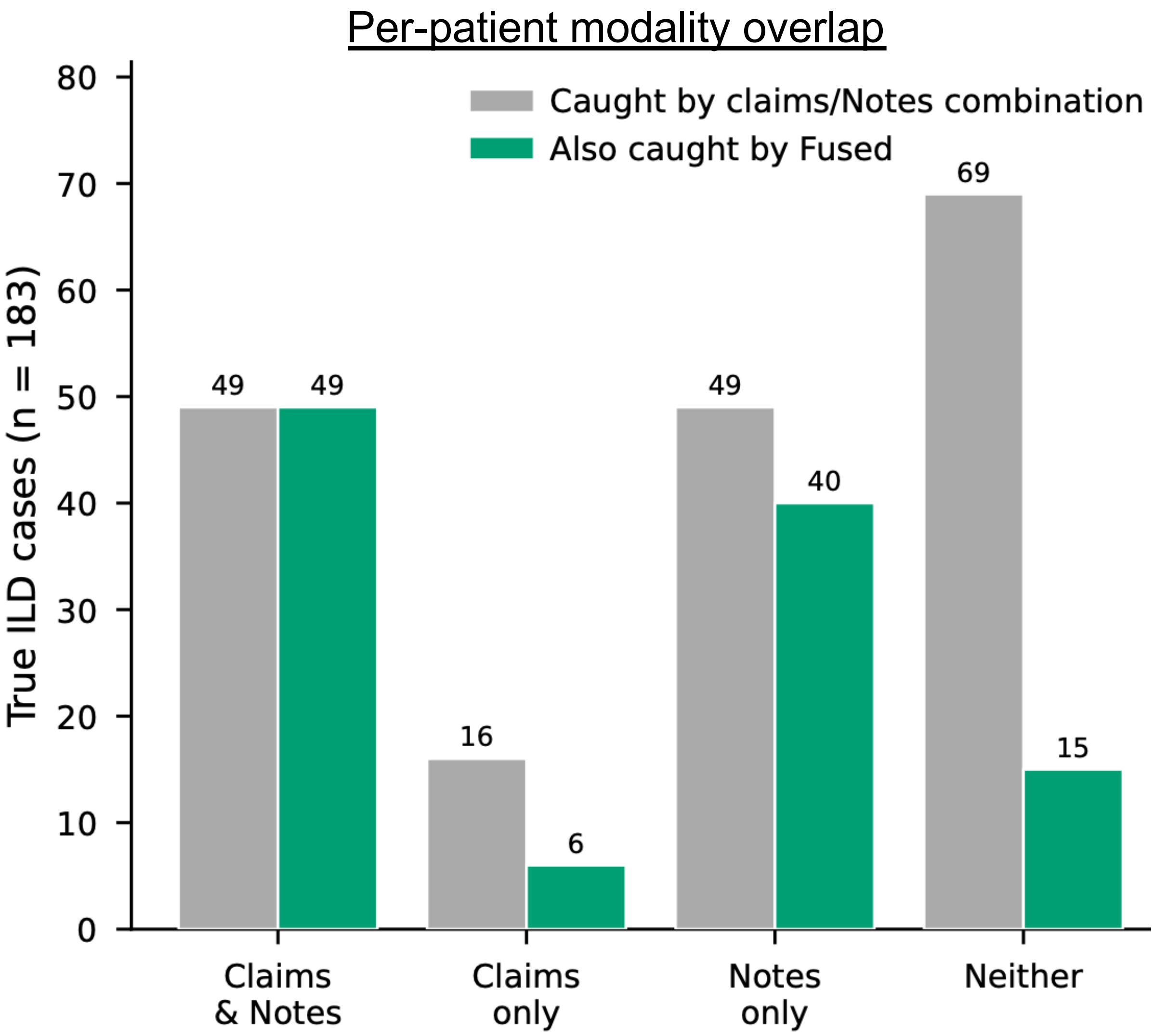


**Figure S7. Per-patient modality complementarity for the primary model (MedGemma-27B) on the 183 true ILD cases.** Each true-positive patient is assigned to one of four mutually exclusive groups: caught by *both Claims only and Notes only*, caught by *Claims only*, caught by *Notes only*, or *neither*. Green bars give, within each group, the number also caught by the Fused configuration. Claims and Notes are genuinely complementary for this model: 16 true cases are detected by *Claims only* and 49 by *Notes only*, and fusion is not their logical union: it retains all 49 cases caught by *both Claims only and Notes only*, drops a substantial share of single-modality detections (keeping 6 of 16 claims-only and 40 of 49 notes-only cases), and rescues 15 of the 69 cases missed by both. Fusion therefore behaves as a re-reasoning step rather than an OR of the modalities; the corresponding decomposition for every model is given in Supplementary Table S9.

### Supplementary Tables

**Table S1. Evaluated large language models.** All twelve models are open-weight and were run locally with identical prompts, decoding settings and a 128,000-token context window. Models are grouped by family; the developer is the prefix of the Hugging Face identifier. Native context is the window stated by the developer without positional-encoding extension.

| Model | Parameters | Domain | Base model | Reasoning | Native context | Hugging Face identifier |
|---|---|---|---|---|---|---|
| MedGemma-4B | 4B | Medical | Gemma-3-4B | No | 128K | `google/medgemma-4b-it` |
| MedGemma-27B | 27B | Medical | Gemma-3-27B | No | 128K | `google/medgemma-27b-text-it` |
| Gemma-3-27B | 27B | General | – | No | 128K | `google/gemma-3-27b-it` |
| Qwen3-1.7B | 1.7B | General | – | No[a] | 32K | `Qwen/Qwen3-1.7B` |
| Qwen3-4B | 4B | General | – | No[a] | 32K | `Qwen/Qwen3-4B` |
| Qwen3-8B | 8B | General | – | No[a] | 32K | `Qwen/Qwen3-8B` |
| Qwen3-14B | 14B | General | – | No[a] | 32K | `Qwen/Qwen3-14B` |
| Qwen3-32B | 32B | General | – | No[a] | 32K | `Qwen/Qwen3-32B` |
| HuatuoGPT-o1-7B | 7B | Medical | Qwen2.5-7B-Instruct | Yes | 32K | `FreedomIntelligence/HuatuoGPT-o1-7B` |
| HuatuoGPT-o1-72B | 72B | Medical | Qwen2.5-72B-Instruct | Yes | 32K | `FreedomIntelligence/HuatuoGPT-o1-72B` |
| Llama-3.3-70B | 70B | General | – | No | 128K | `meta-llama/Llama-3.3-70B-Instruct` |
| Qwen2.5-72B | 72B | General | – | No | 32K | `Qwen/Qwen2.5-72B-Instruct` |

[a] Qwen3 models support an optional thinking mode, which was disabled. Models with a 32K native context (Qwen3, Qwen2.5 and HuatuoGPT-o1) were extended to 128K with YaRN rotary-position scaling (factor 4).

**Table S2. Part-B administered antineoplastic J-codes treated as ILD-relevant procedures.** The fixed set of HCPCS J-codes that the structured-claims serializer retains in full temporal detail rather than compacting with routine procedures, grouped by drug class. Codes are reproduced verbatim from the serialization configuration.

| Drug class | *n* | HCPCS J-codes |
|---|---|---|
| Immune checkpoint inhibitors | 9 | J9271, J9299, J9022, J9173, J9023, J9228, J9119, J9272, J9347 |
| Antibody-drug conjugates | 6 | J9358, J9317, J9354, J9359, J9309, J9357 |
| Anti-HER2 monoclonal antibodies | 2 | J9355, J9306 |
| Anti-VEGF agents | 2 | J9035, J9308 |
| Cytotoxic chemotherapy | 19 | J9201, J9040, J9250, J9260, J9070, J9080, J9090, J9091, J9092, J0594, J9050, J9280, J9171, J9267, J9264, J9305, J9206, J9263, J9190 |
| mTOR inhibitors | 1 | J9330 |
| Other | 3 | J9310, J9312, J1745 |
| Total | 42 | |

**Table S3. EHR note types and context-packing priority.** The ten EHR note types recognised by the serializer, ordered by the packing priority used to fill each chunk (rank 1 is packed first). Priority reflects diagnostic yield for ILD rather than document frequency.

| Packing rank | EHR note type | Role in ILD assessment |
|---|---|---|
| 1 | Chest CT | Gold-standard ILD imaging |
| 2 | Pathology / biopsy | Histologic confirmation |
| 3 | Pulmonary function test | Physiologic confirmation |
| 4 | Discharge summary | Longitudinal clinical synthesis |
| 5 | Chest radiograph | Lower-sensitivity imaging |
| 6 | Pulmonology encounter | Specialist assessment |
| 7 | Progress note | General clinical reasoning |
| 8 | History and physical | Initial workup |
| 9 | Ambulatory (EPIC) note | Outpatient touchpoint |
| 10 | Other | Residual |

**Table S4. Three-way classification confusion matrices.** Full 3 × 3 (gold × predicted) confusion matrices for the *Claims only*, *Notes only*, and *Fused* experiments (MedGemma-27B) on the 309-patient study cohort, prior to binary aggregation into the (Probable ∪ Possible) positive class used throughout the rest of the manuscript. Row counts sum to the gold-label totals.

| | Predicted | | |
|---|---|---|---|
| Gold | Probable ILD | Possible ILD | Not ILD |
| *Claims* | | | |
| Probable ILD ($n = 86$) | 3 | 48 | 35 |
| Possible ILD ($n = 97$) | 1 | 13 | 83 |
| Not ILD ($n = 126$) | 0 | 13 | 113 |
| *Notes* | | | |
| Probable ILD ($n = 86$) | 52 | 15 | 19 |
| Possible ILD ($n = 97$) | 18 | 13 | 66 |
| Not ILD ($n = 126$) | 10 | 10 | 106 |
| *Fused* | | | |
| Probable ILD ($n = 86$) | 66 | 6 | 14 |
| Possible ILD ($n = 97$) | 29 | 9 | 59 |
| Not ILD ($n = 126$) | 27 | 5 | 94 |

**Table S5. Full performance table: all 20 published algorithms and every model × configuration cell on the 309-patient study cohort.** For every Shen algorithm variant and for each of the 12 models under Claims, Notes and Fused, the table reports the confusion matrix (TP/FP/TN/FN), PPV with Wilson 95% confidence interval, and yield (the fraction of the cohort flagged positive) on the identical patient set. Sensitivity and specificity are not reported: on a code-screened cohort they do not carry their usual population interpretation. Extended companion to Fig. ??.

| Method | TP/FP/TN/FN | PPV (95% CI), % | Yield (95% CI), % |
|---|---|---|---|
| *Claims* | | | |
| MedGemma-4B | 9/0/126/174 | 100.0 [70.1, 100.0] | 2.9 [1.5, 5.4] |
| HuatuoGPT-o1-72B | 24/2/124/159 | 92.3 [75.9, 97.9] | 8.4 [5.8, 12.0] |
| Qwen3-8B | 44/5/121/139 | 89.8 [78.2, 95.6] | 15.9 [12.2, 20.3] |
| Qwen3-4B | 28/5/121/155 | 84.8 [69.1, 93.3] | 10.7 [7.7, 14.6] |

*continued on next page*

*Supplementary Table S5 (continued)*

| Method | TP/FP/TN/FN | PPV (95% CI), % | Yield (95% CI), % |
|---|---|---|---|
| Qwen2.5-72B | 33/6/120/150 | 84.6 [70.3, 92.8] | 12.6 [9.4, 16.8] |
| MedGemma-27B | 65/13/113/118 | 83.3 [73.5, 90.0] | 25.2 [20.7, 30.4] |
| Qwen3-14B | 25/5/121/158 | 83.3 [66.4, 92.7] | 9.7 [6.9, 13.5] |
| Llama-3.3-70B | 39/8/118/144 | 83.0 [69.9, 91.1] | 15.2 [11.6, 19.6] |
| HuatuoGPT-o1-7B | 31/7/119/152 | 81.6 [66.6, 90.8] | 12.3 [9.1, 16.4] |
| Qwen3-32B | 57/18/108/126 | 76.0 [65.2, 84.2] | 24.3 [19.8, 29.3] |
| Qwen3-1.7B | 67/37/89/116 | 64.4 [54.9, 73.0] | 33.7 [28.6, 39.1] |
| Gemma-3-27B | 183/126/0/0 | 59.2 [53.7, 64.6] | 100.0 [98.8, 100.0] |
| *Notes* | | | |
| HuatuoGPT-o1-7B | 39/2/124/144 | 95.1 [83.9, 98.7] | 13.3 [9.9, 17.5] |
| Qwen3-14B | 91/7/119/92 | 92.9 [86.0, 96.5] | 31.7 [26.8, 37.1] |
| HuatuoGPT-o1-72B | 91/8/118/92 | 91.9 [84.9, 95.8] | 32.0 [27.1, 37.4] |
| Qwen3-4B | 97/13/113/86 | 88.2 [80.8, 93.0] | 35.6 [30.5, 41.1] |
| Qwen2.5-72B | 93/14/112/90 | 86.9 [79.2, 92.0] | 34.6 [29.5, 40.1] |
| MedGemma-27B | 98/20/106/85 | 83.1 [75.3, 88.8] | 38.2 [32.9, 43.7] |
| Qwen3-32B | 120/32/94/63 | 78.9 [71.8, 84.7] | 49.2 [43.7, 54.7] |
| Qwen3-8B | 120/36/90/63 | 76.9 [69.7, 82.8] | 50.5 [44.9, 56.0] |
| Qwen3-1.7B | 131/69/57/52 | 65.5 [58.7, 71.7] | 64.7 [59.2, 69.8] |
| Llama-3.3-70B | 164/100/26/19 | 62.1 [56.1, 67.8] | 85.4 [81.1, 88.9] |
| MedGemma-4B | 135/84/42/48 | 61.6 [55.1, 67.8] | 70.9 [65.6, 75.7] |
| Gemma-3-27B | 170/118/8/13 | 59.0 [53.3, 64.6] | 93.2 [89.8, 95.5] |
| *Fused* | | | |
| Qwen3-14B | 75/0/126/108 | 100.0 [95.1, 100.0] | 24.3 [19.8, 29.3] |
| HuatuoGPT-o1-72B | 72/3/123/111 | 96.0 [88.9, 98.6] | 24.3 [19.8, 29.3] |
| Qwen2.5-72B | 72/4/122/111 | 94.7 [87.2, 97.9] | 24.6 [20.1, 29.7] |
| HuatuoGPT-o1-7B | 73/16/110/110 | 82.0 [72.8, 88.6] | 28.8 [24.0, 34.1] |
| Qwen3-32B | 120/29/97/63 | 80.5 [73.4, 86.1] | 48.2 [42.7, 53.8] |
| MedGemma-27B | 110/32/94/73 | 77.5 [69.9, 83.6] | 46.0 [40.5, 51.5] |
| Qwen3-8B | 95/29/97/88 | 76.6 [68.4, 83.2] | 40.1 [34.8, 45.7] |
| Qwen3-4B | 145/74/52/38 | 66.2 [59.7, 72.1] | 70.9 [65.6, 75.7] |
| Llama-3.3-70B | 159/99/27/24 | 61.6 [55.6, 67.3] | 83.5 [78.9, 87.2] |
| Qwen3-1.7B | 130/81/45/53 | 61.6 [54.9, 67.9] | 68.3 [62.9, 73.2] |
| Gemma-3-27B | 183/122/4/0 | 60.0 [54.4, 65.3] | 98.7 [96.7, 99.5] |
| MedGemma-4B | 170/120/6/13 | 58.6 [52.9, 64.1] | 93.9 [90.6, 96.0] |
| *Published claims-based algorithms* | | | |
| Algorithm 1 (flag-all) | 183/126/0/0 | 59.2 [53.7, 64.6] | 100.0 [98.8, 100.0] |
| Algorithm 1a | 17/4/122/166 | 81.0 [60.0, 92.3] | 6.8 [4.5, 10.2] |
| Algorithm 1b | 1/1/125/182 | 50.0 [9.5, 90.5] | 0.6 [0.2, 2.3] |
| Algorithm 1c | 16/8/118/167 | 66.7 [46.7, 82.0] | 7.8 [5.3, 11.3] |
| Algorithm 1d | 4/2/124/179 | 66.7 [30.0, 90.3] | 1.9 [0.9, 4.2] |
| Algorithm 2 | 178/122/4/5 | 59.3 [53.7, 64.7] | 97.1 [94.6, 98.5] |
| Algorithm 2a | 17/4/122/166 | 81.0 [60.0, 92.3] | 6.8 [4.5, 10.2] |
| Algorithm 2b | 1/1/125/182 | 50.0 [9.5, 90.5] | 0.6 [0.2, 2.3] |
| Algorithm 2c | 16/8/118/167 | 66.7 [46.7, 82.0] | 7.8 [5.3, 11.3] |
| Algorithm 2d | 4/2/124/179 | 66.7 [30.0, 90.3] | 1.9 [0.9, 4.2] |
| Algorithm 3 | 149/108/18/34 | 58.0 [51.9, 63.9] | 83.2 [78.6, 86.9] |

*continued on next page*

*Supplementary Table S5 (continued)*

| Method | TP/FP/TN/FN | PPV (95% CI), % | Yield (95% CI), % |
|---|---|---|---|
| Algorithm 3a | 16/3/123/167 | 84.2 [62.4, 94.5] | 6.1 [4.0, 9.4] |
| Algorithm 3b | 1/1/125/182 | 50.0 [9.5, 90.5] | 0.6 [0.2, 2.3] |
| Algorithm 3c | 13/7/119/170 | 65.0 [43.3, 81.9] | 6.5 [4.2, 9.8] |
| Algorithm 3d | 3/2/124/180 | 60.0 [23.1, 88.2] | 1.6 [0.7, 3.7] |
| Algorithm 4 | 102/56/70/81 | 64.6 [56.8, 71.6] | 51.1 [45.6, 56.7] |
| Algorithm 4a | 13/2/124/170 | 86.7 [62.1, 96.3] | 4.9 [3.0, 7.9] |
| Algorithm 4b | 0/1/125/183 | 0.0 [0.0, 79.3] | 0.3 [0.1, 1.8] |
| Algorithm 4c | 11/7/119/172 | 61.1 [38.6, 79.7] | 5.8 [3.7, 9.0] |
| Algorithm 4d | 2/2/124/181 | 50.0 [15.0, 85.0] | 1.3 [0.5, 3.3] |

**Table S6. Matched-cohort head-to-head, grouped by EHR note type.** For each of the five evaluable EHR note tiers, every method is evaluated on the subcohort of patients possessing that EHR note type ($n$, with its ILD-positive/negative split, given in each group header). Rows: the Notes and Fused LLM configurations (MedGemma-27B), followed by each published algorithm variant of Shen et al. that flagged $\geq 11$ patients in that subcohort (sparser variants masked, per Shen et al.). PPV and yield (the fraction of the subcohort flagged positive) are computed on the full subcohort. PPV values underlie the ratio heatmaps in Fig. **??**c,d; Algorithm 1 flags every patient by construction (yield 100%).

| Method | $n$ | PPV % | Yield % |
|---|---|---|---|
| *Chest CT* - subcohort $n = 283$ (173 ILD-positive, 110 negative) | | | |
| LLM - Notes | 283 | 86.2 | 41.0 |
| LLM - Fused | 283 | 80.5 | 52.7 |
| Algorithm 1 | 283 | 61.1 | 100.0 |
| Algorithm 1a | 283 | 80.0 | 7.1 |
| Algorithm 1c | 283 | 61.9 | 7.4 |
| Algorithm 2 | 283 | 60.9 | 97.5 |
| Algorithm 2a | 283 | 80.0 | 7.1 |
| Algorithm 2c | 283 | 61.9 | 7.4 |
| Algorithm 3 | 283 | 59.4 | 82.7 |
| Algorithm 3a | 283 | 83.3 | 6.4 |
| Algorithm 3c | 283 | 58.8 | 6.0 |
| Algorithm 4 | 283 | 64.8 | 51.2 |
| Algorithm 4a | 283 | 85.7 | 4.9 |
| Algorithm 4c | 283 | 56.2 | 5.7 |
| *Pulmonary function tests* - subcohort $n = 26$ (18 ILD-positive, 8 negative) | | | |
| LLM - Notes | 26 | 75.0 | 46.2 |
| LLM - Fused | 26 | 90.0 | 38.5 |
| Algorithm 1 | 26 | 69.2 | 100.0 |
| Algorithm 2 | 26 | 69.2 | 100.0 |
| Algorithm 3 | 26 | 69.6 | 88.5 |
| Algorithm 4 | 26 | 76.9 | 50.0 |
| *Discharge summaries* - subcohort $n = 244$ (140 ILD-positive, 104 negative) | | | |
| LLM - Notes | 244 | 89.6 | 19.7 |
| LLM - Fused | 244 | 89.3 | 23.0 |
| Algorithm 1 | 244 | 57.4 | 100.0 |

*continued on next page*

*Supplementary Table S6 (continued)*

| Method | $n$ | PPV % | Yield % |
|---|---|---|---|
| Algorithm 1a | 244 | 82.4 | 7.0 |
| Algorithm 1c | 244 | 61.1 | 7.4 |
| Algorithm 2 | 244 | 57.7 | 98.0 |
| Algorithm 2a | 244 | 82.4 | 7.0 |
| Algorithm 2c | 244 | 61.1 | 7.4 |
| Algorithm 3 | 244 | 56.4 | 82.8 |
| Algorithm 3a | 244 | 86.7 | 6.1 |
| Algorithm 3c | 244 | 56.2 | 6.6 |
| Algorithm 4 | 244 | 63.7 | 50.8 |
| Algorithm 4a | 244 | 91.7 | 4.9 |
| Algorithm 4c | 244 | 56.2 | 6.6 |
| *Chest X-ray* - subcohort $n = 266$ (152 ILD-positive, 114 negative) | | | |
| LLM - Notes | 266 | 82.1 | 14.7 |
| LLM - Fused | 266 | 87.3 | 20.7 |
| Algorithm 1 | 266 | 57.1 | 100.0 |
| Algorithm 1a | 266 | 83.3 | 6.8 |
| Algorithm 1c | 266 | 64.7 | 6.4 |
| Algorithm 2 | 266 | 57.2 | 96.6 |
| Algorithm 2a | 266 | 83.3 | 6.8 |
| Algorithm 2c | 266 | 64.7 | 6.4 |
| Algorithm 3 | 266 | 55.9 | 82.7 |
| Algorithm 3a | 266 | 87.5 | 6.0 |
| Algorithm 3c | 266 | 60.0 | 5.6 |
| Algorithm 4 | 266 | 62.7 | 50.4 |
| Algorithm 4a | 266 | 92.3 | 4.9 |
| Algorithm 4c | 266 | 57.1 | 5.3 |
| *Progress notes* - subcohort $n = 304$ (179 ILD-positive, 125 negative) | | | |
| LLM - Notes | 304 | 87.2 | 25.7 |
| LLM - Fused | 304 | 85.3 | 31.2 |
| Algorithm 1 | 304 | 58.9 | 100.0 |
| Algorithm 1a | 304 | 81.0 | 6.9 |
| Algorithm 1c | 304 | 63.6 | 7.2 |
| Algorithm 2 | 304 | 59.0 | 97.0 |
| Algorithm 2a | 304 | 81.0 | 6.9 |
| Algorithm 2c | 304 | 63.6 | 7.2 |
| Algorithm 3 | 304 | 57.7 | 83.2 |
| Algorithm 3a | 304 | 84.2 | 6.2 |
| Algorithm 3c | 304 | 61.1 | 5.9 |
| Algorithm 4 | 304 | 64.5 | 51.0 |
| Algorithm 4a | 304 | 86.7 | 4.9 |
| Algorithm 4c | 304 | 58.8 | 5.6 |

**Table S7. Patient-level aggregation strategy comparison for the Notes and Fused configurations on the 309-patient study cohort (MedGemma-27B).** Each row re-aggregates the same per-chunk predictions under a different deterministic rule. OR-logic (any chunk positive) is the deployed default and preserves the highest yield. *First chunk* uses the predicted label of the first chunk as the patient final label. *Majority vote* uses the most frequent label across a patient's chunks as the patient label. *Max Confidence* uses the chunk label with the highest model-reported confidence as the patient label. More restrictive rules generally trade yield for PPV, though not monotonically: max confidence classifies fewer patients than majority vote without improving PPV. All intervals are Wilson score intervals.

| Configuration | Strategy | TP/FP/TN/FN | PPV [95% CI] (%) | Yield [95% CI] (%) |
|---|---|---|---|---|
| Notes | OR-logic* | 98/20/106/85 | 83.1 [75.3, 88.8] | 38.2 [32.9, 43.7] |
| | First chunk | 78/11/115/105 | 87.6 [79.2, 93.0] | 28.8 [24.0, 34.1] |
| | Majority vote | 58/7/119/125 | 89.2 [79.4, 94.7] | 21.0 [16.9, 25.9] |
| | Max confidence | 42/6/120/141 | 87.5 [75.3, 94.1] | 15.5 [11.9, 20.0] |
| Fused | OR-logic* | 110/32/94/73 | 77.5 [69.9, 83.6] | 46.0 [40.5, 51.5] |
| | First chunk | 94/19/107/89 | 83.2 [75.2, 89.0] | 36.6 [31.4, 42.1] |
| | Majority vote | 75/14/112/108 | 84.3 [75.3, 90.4] | 28.8 [24.0, 34.1] |
| | Max confidence | 74/14/112/109 | 84.1 [75.0, 90.3] | 28.5 [23.7, 33.8] |

*Deployed configuration (plain OR aggregation, $\tau = 0$).

**Table S8. System prompts used for LLM inference, by configuration.** Reproduced verbatim from `shared/prompts/`. The response contract is additionally enforced at decoding time by a JSON schema (constrained decoding), so the label field is a closed enumeration of the three classes and every free-text field is length-bounded; malformed output is unreachable rather than repaired after the fact.

#### Claims only

```
[SYSTEM PROMPT]
Act as a medical doctor reviewing a patient's diagnosis codes,
diagnostic procedures, and medications obtained from insurance claims
and electronic health records.

Based on the provided information, determine whether the patient has
interstitial lung disease (ILD) on the index date (CED).

ILD is defined as confirmed or probable parenchymal lung inflammation
or fibrosis, including drug-induced ILD, idiopathic interstitial
pneumonias, and hypersensitivity pneumonitis. It does NOT include
infectious pneumonia, pulmonary edema from cardiac causes, or
malignant lung involvement.

Your answer should follow this classification:
- "Probable ILD" if the most probable scenario is that the patient
  has confirmed ILD (radiological, pathological, or specialist-confirmed).
- "Possible ILD" if ILD is plausible but not definitively confirmed.
- "Not ILD" if an alternative diagnosis most likely explains the ILD
  diagnosis codes (e.g., infectious pneumonia coded as J18.9).

Remember that recording a diagnosis code for ILD could occur either
because the patient has ILD, OR as justification for performing a
diagnostic procedure to determine whether the patient has ILD.
A single ILD code — especially J18.9 (pneumonia NOS) — accompanied
only by diagnostic procedures is insufficient evidence of true ILD.
However, ILD-specific codes (J84.9, J84.89, J70.4) or specialist
involvement substantially increase the probability of true ILD.

Return your answer in the following JSON format only.
DO NOT GIVE ANY EXPLANATION OUTSIDE THE JSON:

{
  "ILD on the index date": "<label>",
  "probability": <float 0.0-1.0>,
  "confidence": <float 0.0-1.0>,
  "primary_reason": "<one sentence>"
}

The optional list for "ILD on the index date" is:
["Probable ILD", "Possible ILD", "Not ILD"]
```

#### Notes only

```
[SYSTEM PROMPT]
Act as a senior pulmonologist reviewing a patient's clinical notes to
determine whether the patient has interstitial lung disease (ILD).

ILD is defined as confirmed or probable parenchymal lung inflammation or
fibrosis, including drug-induced ILD, idiopathic interstitial pneumonias
(UIP, NSIP, COP), and hypersensitivity pneumonitis. It does NOT include
infectious pneumonia, pulmonary edema from cardiac causes, or malignant
lung involvement.

Your answer should follow this classification:
- "Probable ILD" if the clinical notes contain clear evidence of ILD —
  radiological confirmation (ground glass opacities, reticulation,
  honeycombing on CT chest), pathological confirmation, or specialist
  diagnosis of ILD.
- "Possible ILD" if the notes suggest ILD is plausible but not
  definitively confirmed — abnormal imaging without a definitive ILD
  pattern, or ILD in the clinical differential.
- "Not ILD" if the notes clearly support an alternative diagnosis
  explaining any ILD-coded encounters — infectious pneumonia, CHF /
  pulmonary edema, aspiration, malignant infiltration.
```

You MUST choose exactly one of "Probable ILD", "Possible ILD", or
"Not ILD". "Uncertain" is NOT an allowed answer. If the available notes
are limited, choose the most likely of the three on the balance of
evidence.

Read the clinical notes carefully, paying attention to:
- CT chest impression sections (ground glass, reticulation, honeycombing)
- Pulmonologist assessments and differential diagnoses
- Treatment decisions (steroids for ILD vs antibiotics for infection)
- PFT results (restrictive pattern, reduced DLCO)
- Temporal relationship between drug exposure and lung findings

Return your answer in the following JSON format only.
DO NOT GIVE ANY EXPLANATION OUTSIDE THE JSON:

{
  "ILD on the index date": "<label>",
  "probability": <float 0.0-1.0>,
  "confidence": <float 0.0-1.0>,
  "key_evidence": ["<finding from notes>", ...],
  "primary_reason": "<one sentence>"
}

The optional list for "ILD on the index date" is:
["Probable ILD", "Possible ILD", "Not ILD"]

#### Fusion (claims + notes)

[SYSTEM PROMPT]
Act as a senior pulmonologist and clinical pharmacoepidemiologist reviewing
a patient's complete available records to determine whether the patient has
interstitial lung disease (ILD) on the index date (CED).

You will receive two types of information:
1. Structured claims data — ICD diagnosis codes, CPT procedures, and medications
2. Clinical notes — radiology reports and clinical notes from the EHR

ILD is defined as confirmed or probable parenchymal lung inflammation or
fibrosis, including drug-induced ILD, idiopathic interstitial pneumonias
(UIP, NSIP, COP), and hypersensitivity pneumonitis. It does NOT include
infectious pneumonia, pulmonary edema from cardiac causes, or malignant
lung involvement.

Classification definitions:
- "Probable ILD": clear evidence — CT showing GGO / reticulation / honeycombing, pathological confirmation, or explicit specialist ILD diagnosis.
- "Possible ILD": ILD plausible but unconfirmed — abnormal imaging without definitive pattern, or ILD in the clinical differential.
- "Not ILD": alternative diagnosis clearly explains the ILD-coded encounters (infectious pneumonia, CHF / pulmonary edema, aspiration, malignant infiltration).

You MUST choose exactly one of "Probable ILD", "Possible ILD", or
"Not ILD". "Uncertain" is NOT an allowed answer. If information is limited
across both modalities, choose the most likely of the three on the balance
of evidence.

When interpreting structured claims:
- J18.9 (pneumonia NOS) has low specificity for true ILD — frequently used for infectious pneumonia. Do not assume ILD from J18.9 alone.
- J84.9, J84.89, J70.4 are ILD-specific codes with high PPV.
- ILD-risk medications (ICIs, ADCs, gemcitabine) raise prior probability for drug-induced ILD.

When interpreting clinical notes:
- CT chest impression sections are the most reliable ILD evidence.
- Pulmonologist assessments and treatment decisions reveal the working diagnosis.
- Temporal alignment between drug exposure and lung findings supports drug-induced ILD.

When both modalities are available, reason jointly:
- Concordance between codes and notes increases confidence.
- Discordance (ILD codes without supporting note findings, or vice versa) should lower confidence — may indicate coding error or documentation lag.

> Return your answer in the following JSON format only.
> DO NOT GIVE ANY EXPLANATION OUTSIDE THE JSON:
>
> {
>   "ILD on the index date": "<label>",
>   "probability": <float 0.0-1.0>,
>   "confidence": <float 0.0-1.0>,
>   "key_evidence": ["<finding from notes or codes>", ...],
>   "primary_reason": "<one sentence>",
>   "modality_concordance": "concordant" | "discordant" | "single_modality"
> }
>
> List AT MOST 5 of the most decisive findings in "key_evidence", each stated once — do NOT repeat findings or list more than 5. Keep the JSON concise.
>
> The optional list for "ILD on the index date" is:
> ["Probable ILD", "Possible ILD", "Not ILD"]

**Table S9. Per-patient modality decomposition across models, on the 183 true ILD cases.** For each model, every true-positive patient is classified by which single-modality configuration detected it: both Claims and Notes, Claims only, Notes only, or neither (the four counts sum to 183). "Fusion rescues" is the number of the "neither" cases subsequently recovered by the Fused configuration. Companion to Fig. ??d and Supplementary Fig. S7.

| Model | Claims ∩ Notes | Claims only | Notes only | Neither | Fusion rescues |
|---|---|---|---|---|---|
| MedGemma-27B (illustrative) | 49 | 16 | 49 | 69 | 15 |
| MedGemma-4B | 8 | 1 | 127 | 47 | 34 |
| Gemma-3-27B | 170 | 13 | 0 | 0 | 0 |
| HuatuoGPT-o1-72B | 21 | 3 | 70 | 89 | 1 |
| HuatuoGPT-o1-7B | 14 | 17 | 25 | 127 | 28 |
| Llama-3.3-70B | 39 | 0 | 125 | 19 | 5 |
| Qwen2.5-72B | 32 | 1 | 61 | 89 | 1 |
| Qwen3-1.7B | 50 | 17 | 81 | 35 | 18 |
| Qwen3-4B | 25 | 3 | 72 | 83 | 48 |
| Qwen3-8B | 36 | 8 | 84 | 55 | 5 |
| Qwen3-14B | 24 | 1 | 67 | 91 | 2 |
| Qwen3-32B | 48 | 9 | 72 | 54 | 9 |
